# A randomized double-blind Phase IIb trial to evaluate the efficacy of ChAd63-KH for the treatment of post kala-azar dermal leishmaniasis

**DOI:** 10.1101/2024.04.11.24305597

**Authors:** Brima M. Younis, Rebecca Wiggins, Eltahir A.G. Khalil, Mohamed Osman, Francesco Santoro, Chiara Sonnati, Ada Keding, Maria Novedrati, Giorgio Montesi, Ali Noureldein, Elmukashfi T. A. Elmukashfi, Ala Eldin Mustafa, Mohammed Alamin, Mohammed Saeed, Khalid Salman, Ahmed J. Suliman, Amin E.A. Musa, Alison M. Layton, Charles J. N. Lacey, Paul M. Kaye, Ahmed M. Musa

## Abstract

**Background:** In a recent Phase IIa clinical trial, the candidate leishmaniasis vaccine ChAd63-KH was shown to be safe and immunogenic in Sudanese patients with post kala-azar dermal leishmaniasis (PKDL). However, its value as a stand-alone therapeutic was unknown.

**Methods:** To assess the therapeutic efficacy of ChAd63-KH, we conducted a “window of opportunity” randomized, double-blind, placebo-controlled trial (Clinicaltrials.gov registration: NCT03969134). We aimed to enrol 100 participants (male and female aged 12-50 years) with uncomplicated PKDL of ≥ six months duration. ChAd63-KH (7.5×10^10^ viral particles) or saline placebo was administered once intramuscularly. Primary outcomes were safety and efficacy. Safety was determined by adverse event monitoring. Efficacy was the proportion of participants at 90 days post-vaccination with ≥90% improvement in clinical disease. Participants failing to reach this clinical endpoint were offered a standard of care (AmBisome^®^). Secondary outcomes included changes in PKDL severity grade and measurements of vaccine-induced immune response.

**Findings:** Between 4^th^ April 2020 and 17^th^ June 2022, 86 participants (66 adolescents, 20 adults; 47% female, 53% male) were enrolled and randomised to receive ChAd63-KH or placebo. 75 participants (87%) completed the trial as per protocol. No severe or serious adverse events were observed. At day 90 post vaccination, 6/40 (15%) and 4/35 (11%) participants in the vaccine and placebo groups respectively showed ≥ 90% clinical improvement (RR 1.31 [95% CI, 0.40 to 4.28], p=0.742). There were also no significant differences in PKDL grade between study arms. Whole blood transcriptomic analysis identified transcriptional modules associated with interferon responses and monocyte and dendritic cell activation, confirming vaccine reactogenicity.

**Interpretation:** Single dose administration of ChAd63-KH vaccine had no therapeutic efficacy in this subset of Sudanese PKDL patients. Further studies are needed to evaluate whether this vaccine would have therapeutic benefit using alternate dosing regimens or in combination with standard chemotherapy or immune modulation, and whether it has efficacy as a prophylactic vaccine for cutaneous or visceral leishmaniasis.

**Funding:** This study was funded by the Wellcome Trust.

**Research in context:** *Evidence before this study:* A leishmaniasis vaccine candidate was developed employing chimpanzee adenovirus 63 (ChAd63) to deliver genes encoding two *Leishmania* antigens, KMP-11 and HASPB1. This vaccine (ChAd63-KH) was previously evaluated for safety and immunogenicity in a Phase I healthy volunteer study and a Phase IIa study in Sudanese patients with post kala-azar dermal leishmaniasis (PKDL). It was shown to be safe and immunogenic, warranting further clinical studies to evaluate efficacy as a stand-alone therapeutic in PKDL patients.

*Added value of this study:* This clinical trial was designed to evaluate the safety and efficacy of ChAd63-KH in PKDL patients with persistent disease (dermal lesions for ≥ 6 months). If successful, single dose vaccination would significantly improve treatment options currently available to patients. The safety of ChAd63-KH was confirmed, with no severe or serious adverse events observed in trial participants. Approximately 13% of participants had ≥90% improvement in their PKDL over the course of 90 days follow up post vaccination, but this did not differ between vaccine and placebo arms, indicating that this reflected spontaneous cure rather than vaccine efficacy. Immune monitoring using whole blood transcriptomics confirmed the previously reported ability of this vaccine to induce immune responses in humans.

*Implications of all the available evidence:* This study indicates that as a stand-alone treatment, single dose vaccination with ChAd63-KH was unable to overcome the immune dysfunction that maintains persistent PKDL. A similar “high bar” has also been encountered in therapeutic vaccine trials for other persistent diseases. Given previous success with other forms of immunochemotherapy in PKDL, future therapeutic vaccine studies in PKDL might also benefit from combining ChAd63-KH vaccination with additional chemotherapy or immune modulation. The prophylactic efficacy of this vaccine against different types of leishmaniasis also remains to be evaluated.

## Introduction

Post kala azar dermal leishmaniasis (PKDL) is a chronic dermatological sequela associated with treatment for visceral leishmaniasis (VL; kala azar), but it may occur without a history of VL or even during treatment of VL (when it is known as para kala-azar dermal leishmaniasis). PKDL typically involves the face and later spreads to the extremities and trunk ^1–3^. PKDL has often been confused with leprosy and because of its chronic but not debilitating nature, it is often tolerated by infected people who fail to seek treatment. Case rates for PKDL are intimately linked to the waxing and waning of VL incidence ^4^. The VL elimination campaign in South Asia has served to focus attention on PKDL, as those with the disease have been shown to harbour parasites in their skin and be infectious to the sand fly vector. As such, people with PKDL may serve to maintain infection in inter-epidemic periods thus posing a threat to elimination ^5^. Epidemiological modelling has demonstrated the value of a PKDL vaccine for mitigating this risk in a post-elimination era ^6^. Recent calls to action for VL elimination on the African continent also recognise the need to have effective means to control PKDL ^7^. Drug regimens for PKDL are generally arduous, invasive and have multiple potential side effects ^8,9^. PKDL can also be exacerbated by HIV, and such cases respond poorly to treatment ^10^. New combination therapies have been evaluated and show promise both in reducing the burden of PKDL ^11^ and in treatment ^12^, but there has been a strong and persistent argument for developing additional means of control including therapeutic vaccines ^13–15^.

PKDL has several intriguing features epidemiologically, clinically and immunologically. VL is caused by infection with two species of the protozoan parasite *Leishmania,* namely *L. donovani and L. infantum,* yet PKDL is restricted to infection with *L. donovani* and found only in the Old World ^2^. PKDL is also restricted within the geographical range of *L. donovani*, being found in Sudan and South Asia, but rarely in other countries in East Africa. PKDL typically occurs after treatment whether this be with pentavalent antimonials, amphotericin B, miltefosine and paromomycin/ antimonial combinations ^16^. However, the onset of disease varies geographically, being rapid (weeks to months) in Sudan compared to delayed (often years) in South Asia. Clinical presentation also varies geographically and at different body sites, with various hypotheses, including UVB exposure, being put forward to explain differences in clinical presentation ^17,18^. In Sudan, disease presents mainly as nodules and papules that in ∼80% of cases self-resolve over several months ^8,19^. In the remainder, lesions may persist often for years. In contrast, in South Asia cases can show hypopigmented macular lesions or be polymorphic, with both macular and nodular lesions. Immunological and histopathological differences have also been noted ^17,18,20–23^. Collectively, these features suggest that PKDL is a heterogeneous disease, with implications for the development of new therapeutics.

Therapeutic vaccination against PKDL in Sudan has its historical roots in the use of first-generation vaccines for VL. Khalil and colleagues conducted Phase I/II randomised trials in volunteers with no history of VL and showed that two intradermal doses of autoclaved *L. major* (ALM) administered with Bacille Calmette-Guérin (BCG) was safe, well tolerated and immunogenic, as measured by skin test conversion ^24–26^. However, no efficacy with regards protection against VL was observed. These studies were followed up by evaluation of alum-adjuvanted ALM + BCG, again demonstrating safety and immunogenicity ^27,28^. This latter formulation was subsequently evaluated as a potential therapeutic in PKDL patients with persistent disease. The study enrolled 30 patients randomised to receive Alum-ALM+BCG + sodium stibogluconate (SSG) vs. SSG alone and reported a cure rate at 60 days of 87% in the combined therapy group vs 53% in the SSG alone group (SSG + vaccine efficacy=71%, 95% CI for risk ratio 0.7-1.16)) ^29^. Based on these encouraging results, we evaluated the safety and immunogenicity of ChAd63-KH, a new adenoviral vaccine incorporating two well-documented vaccine candidate antigens that was designed to have potential as a pan-leishmaniasis vaccine ^30^. This open label Phase IIa therapeutic trial in persistent PKDL patients (LEISH2a; ClinicalTrials.gov ID: NCT02894008) was designed as a “window of opportunity” study, where vaccine was administered alone, and standard of care provided as needed after a 90 day follow up period ^31^. The study confirmed that ChAd63-KH was both safe and immunogenic in this patient group, opening the way for the randomised controlled efficacy trial reported here.

## Methods

### Ethics statement

The LEISH2b study (Clinicaltrials.gov ID: NCT03969134) was approved by the Sudan National Medicines and Poisons Board, and the Ethical Review Committees of the Institute of Endemic Diseases, University of Khartoum and the Department of Biology, University of York. LEISH2b was sponsored by the University of York. The study was conducted according to the principles of the current revision of the Declaration of Helsinki 2008 and ICH guidelines for GCP (CPMP/ICH/135/95). All participants provided written informed consent before enrolment. Consent forms are available accompanying the published protocol^32^.

### Study design and participants

LEISH2b was a randomized, double blind, placebo-controlled therapeutic trial designed to evaluate the safety and efficacy of therapeutic vaccination with the investigational vaccine ChAd63-KH. The initial study design allowed for recruitment of 100 participants diagnosed with PKDL aged between 18-50 (adults) or 12-17 (adolescents) and with persistent PKDL of greater than six months duration. Full details of the inclusion and exclusion criteria are provided in the published Protocol ^32^. Participants were recruited from an endemic area in Gedaref state, Sudan and all study procedures were conducted at the Professor El-Hassan’s Centre for Tropical Medicine, Dooka, Sudan. Clinical monitoring of the study was performed under contract by ClinServ (http://www.clinserv.net). An independent Data Safety and Monitoring Board (DSMB) was established and met throughout the study period. The DSMB Charter is available accompanying the published protocol ^32^.

### Eligibility criteria

Full inclusion and exclusion criteria are provided in the Protocol ^32^. Key inclusion criteria included: age 12 to 50 years on the day of screening; females must be unmarried, single, or widowed; willing and able to give written informed consent/assent; uncomplicated PKDL of ≥ six month duration; otherwise good health; negative for malaria on blood smear; *Leishmania* PCR positive on the screening skin biopsy; willing to undergo urinary pregnancy tests (females only). Key exclusion criteria included: mucosal or conjunctival PKDL; treatment for PKDL within 21 days; negative rK39 strip test; receipt of a live attenuated vaccine within 60 days or other vaccine within 14 days of screening; history of allergic disease or reactions to vaccines or their components; history of severe local or general reaction to vaccination; fever ≥ 39.5°C within 48 hours, anaphylaxis, bronchospasm, laryngeal oedema, collapse, convulsions or encephalopathy within 48 hours; pregnancy, less than 12 weeks postpartum, lactating, or willingness/intention to become pregnant during the study and for three months following vaccination (females only); seropositive for hepatitis B surface antigen (HBsAg) or Hepatitis C (antibodies to HCV); tuberculosis, leprosy, or malnutrition (malnutrition in adults defined as a BMI <18.5, and in children and adolescents (eight-17yrs) as a Z score cut-off value of <-2 SD).

### Vaccine and study procedures

As described previously^31^, ChAd63-KH encodes two leishmanial proteins (KMP-11 and HASPB1) and was manufactured to cGMP by Advent Srl. (Pomezia, Italy; lot B0004; 7.5×10^10^ viral particles (vp) per ml). ChAd63-KH was administered as a single dose in 1ml volume intramuscularly into the deltoid muscle. Participants were monitored in hospital for 7 days post vaccination and thereafter as out-patients on days 21, 42, 90 and 120 post vaccination. Recruitment occurred over four rounds, punctuated by a revolution, a pandemic and a coup. Meetings of the independent DSMB were held at the end of each round of recruitment. Participants were evaluated for clinical response at day 42 and day 90. Those with less than 75% improvement at day 90 were offered standard treatment (AmBisome^®^; 2.5mg/kg/day for 20 days). If improvement was between 75-90%, they were offered conservative treatment or AmBisome®, and those with greater than or equal to 90% clinical improvement were deemed to not require further treatment (and scored as clinical cure). Some participants defaulted from scheduled visits and were evaluated and treated at unscheduled visits based on their availability. Decisions to treat and evaluation of PKDL were performed by two clinicians. Clinical grade of PKDL was also recorded before and after vaccination using the 4-point grading system described previously ^33^.

### Randomisation and Blinding

A computer generated (Stata 16) randomisation list was prepared by the trial statistician, allocating participants 1:1 to either the active or placebo injection, using randomly permuted blocks, stratified by age group (adult or adolescent). The randomisation allocation was communicated to pharmacy staff at the study site through pre-prepared, sealed envelopes with only external sequential participant numbers. During the trial, adults were found to be more difficult to recruit, and the initial aim of an equal number of adolescents and adults in the trial was not feasible. However, the number of available randomisation envelopes in the Sudan was limited for each age group. Therefore, once allocations for the adolescent group had been used up, allocations for the adult stratum were used in order of presentation irrespective of age group, thus creating a single, unstratified randomised sample.

The vaccine and placebo injections were prepared in blacked out syringes labelled only with the participant identification number. The participants and clinical investigators (who administered the IMP and conducted the study follow-up) were blinded as to which injection participants received. The trial statistician (who generated the randomisation list), trial coordinator (who prepared sealed envelopes according to the randomisation list), assistant pharmacist (who prepared the IMP) and study nurse (who delivered the IMP to the clinical investigators) were not blind to the treatment allocation.

### Outcomes

Primary outcome measures were i) safety as recorded as adverse events from clinical examination and evaluation of blood biochemistry and haematology, and ii) efficacy, as determined by the proportion of participants reaching ≥90% improvement in clinical disease. Secondary outcomes included trajectory of lesion improvement and measures of vaccine induced immune response. Planned immune analyses included whole blood transcriptomics and analysis of peripheral lymphocyte responses (Ab and IFNγ ELISpot). Peripheral lymphocyte and antibody responses could not be measured due to sample loss resulting from the military conflict in Sudan.

### Statistical analysis

The statistical analysis plan is provided in the Supplementary Data file. A total sample size of 100 (randomly assigned 1:1 to be vaccinated with ChAd63-KH or placebo) was determined to be sufficient to detect an increase of ≥25% in the proportion of participants achieving clinical cure, assuming 90% power, 5% statistical significance, a spontaneous clearance rate of ≤2% and loss to follow-up of ≤5% (Fisher’s exact test). Baseline characteristics were summarised descriptively. Continuous measures are reported as mean, standard deviation, median, and range while categorical data are reported as counts and percentages. Number of local and systemic AEs per participant are presented as median, minimum, maximum and IQR. The median number of AEs per participant (separately for local and systemic events) were compared between groups using the Mann-Whitney U test. Primary efficacy outcome of >90% clinical improvement between the two arms was evaluated using relative risk ratio (RR) with statistical significance determined by Fisher’s Exact Test. Event numbers were too low to allow for an adjusted regression analysis. Comparison of categorical data across groups was analysed using Fisher’s Exact Test. Analyses were performed using Stata v18, Rv4.3.2 or Prism 10 for macOS (v10.1.1; GraphPad).

### Whole blood transcriptomic analysis

Whole blood samples (2.5 ml) were collected into PAXgene tubes immediately prior to vaccination and at 1 day post vaccination. All reagents and equipment for these analyses were supplied by ThermoFisher Scientific and processes carried out per manufacturers’ protocols, unless otherwise stated. Total RNA was extracted using the PAXgene Blood RNA kit (PreAnalytiX, QIAGEN). RNA was quantified using the Qubit® 2.0 Fluorometer with the RNA HS Assay Kit. ∼50 ng of total RNA was used to construct sequencing libraries with the Ion AmpliSeq™ Transcriptome Human Gene Expression Kit. Libraries were barcoded, purified with 2.5 X Agencourt AMPure XP Magnetic Beads (Beckman Coulter) and then quantified using Ion Library TaqMan Quantitation Kit on a QuantStudio 5. Libraries were diluted to a concentration of about 50 pMol and pooled in groups of 8 for sequencing on Ion PI Chips. Chips were loaded using the Ion Chef System and the IonPI Hi-Q Chef Kit. Sequencing was performed on an Ion Proton Sequencer using Ion PI Hi-Q Sequencing 200 Kit.

Differential gene expression analysis was performed using DeSeq2. After count data normalization, differential gene expression analysis was performed using pooled day 0 data from the two study cohorts as the baseline for all contrasts. Enrichment of blood transcription modules at each time point in the different groups was assessed with the tmod R package, using as an input the lists of differentially expressed genes ranked by the p-value after multiple test correction, as computed by DeSeq2. Significance of module enrichment was assessed using the CERNO statistical test (a modification of Fisher’s combined probability test) and corrected for multiple testing using the Benjamini–Hochberg correction.

### Role of Funders

The funders played no part in study design, data collection, analysis, interpretation, writing or decision to publish. All authors had full access to study data and final responsibility for the decision to submit for publication.

## Results

### Site Initiation and recruitment

The LEISH2b study was conducted over approximately four years, punctuated by periods of significant challenge that resulted in temporary trial suspensions. The site initiation visit (SIV), attended by all field site personnel, trial monitors and trial staff from the Universities of Khartoum and York took place in August 2019 in Addis Ababa, due to a popular uprising and deteriorating security situation in Sudan. Recruitment began on 4^th^ April 2020 and continued through June 2020 with 17 participants recruited in this period. Recruitment was then paused due to the SARS-CoV-2 pandemic. The second round of recruitment took place between December 2020 and February 2021, enrolling 28 participants. A third round of recruitment took place in April and May 2021 adding 23 participants. Delayed by a military coup, the final round of recruitment (May to June 2022) added 18 more, bringing the total to 86 participants. Following discussion with the LEISH2b DSMB, a meeting of the Trial Steering Group with the funders was convened. All attendees agreed that recruitment should halt at 86 participants due to logistical difficulties in completing the trial prior to expiry of the IMP and with the onset of further military clashes. 75/86 (87%) of enrolled participants completed the study to day 90 post vaccination, with last patient, last visit on 20th Sept 2022. Missed visits and losses to follow up were due to occupational priorities. A further day 120 visit was arranged to allow follow up of any drug treatment provided at the completion of the study. A summary of study recruitment is provided in the CONSORT diagram (**Figure 1**).

**Figure 1.**
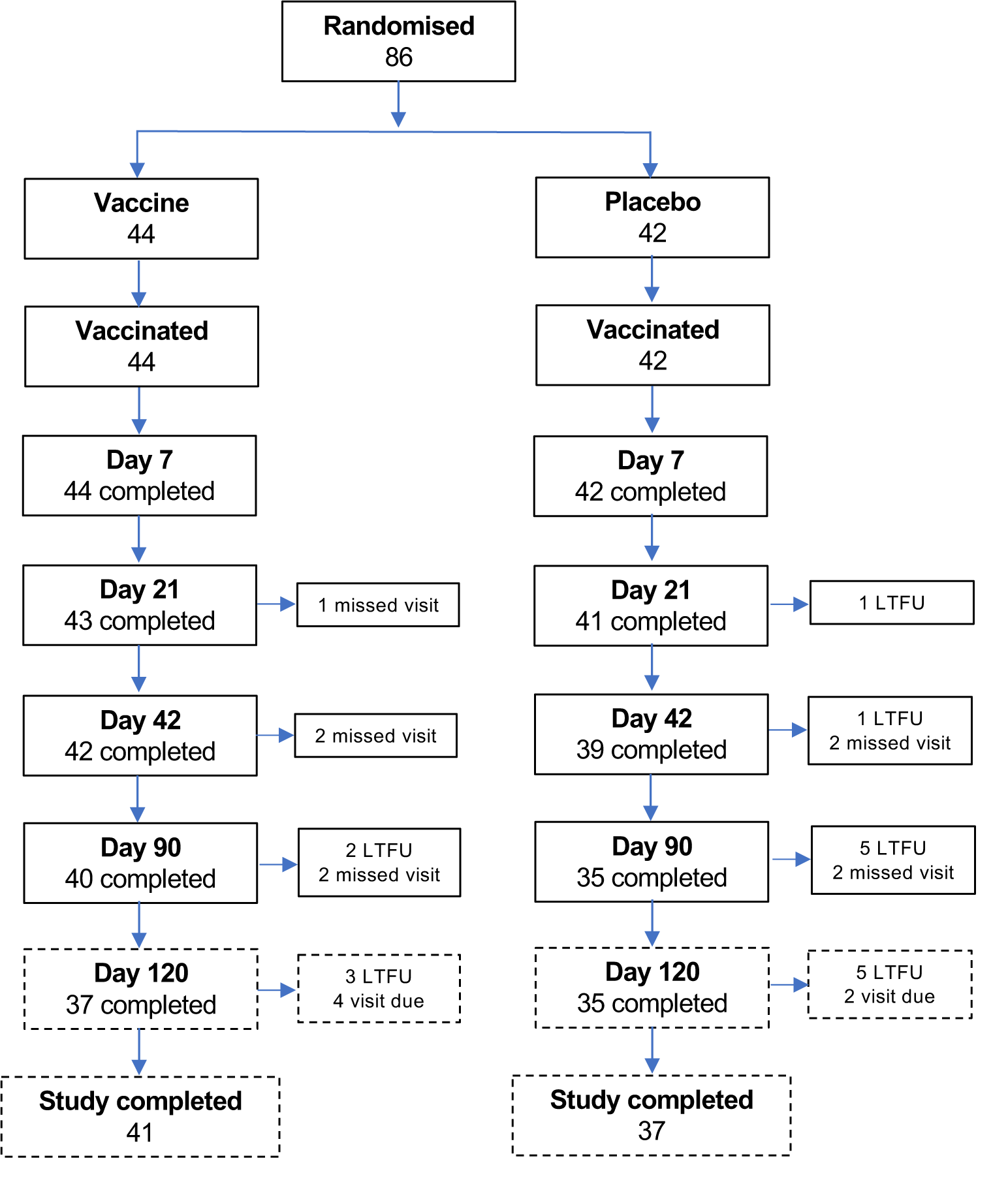
Study CONSORT diagram. Solid boxes indicate participants to primary outcome at day 90 post vaccination. Treatment was offered per protocol from d90 (see text). Dotted boxes were outside trial window for primary outcome and represent scheduled follow to provide or monitor standard of care.

### Study population

Demographics and PKDL grading of the 86 participants enrolled in this study are shown in **Table 1**. All participants had PKDL for 6mths duration or longer with the majority (88%) graded as either PKDL grade 1 or 2 (**Table 1 and Table S1**). Day 90 visits were completed by 75 (87%) participants (n=11missed this time point or had been lost to follow-up; **Figure 1**). As per protocol, participants were offered standard of care (AmBisome®) at day 90 if clinical improvement was less than 90%. Overall, 26/86 (30%) elected to receive treatment (8/20 adults; 18/66 adolescents) and these participants remain in long term follow up to confirm drug effectiveness.

**Table 1.** Baseline characteristics of participants in Leish2b trial.

| Characteristics |  | Vaccine |  | Placebo |  |
| --- | --- | --- | --- | --- | --- |
|  |  | N=44 |  | N=42 |  |
| Adults |  | n=11 |  | n=9 |  |
|  |  | N | % | N | % |
| Sex | Male | 8 | 73 | 6 | 67 |
|  | Female | 3 | 27 | 3 | 33 |
| PKDL grade | Grade 1 | 7 | 64 | 2 | 22 |
|  | Grade 2 | 3 | 27 | 5 | 56 |
|  | Grade 3 | 1 | 9 | 2 | 22 |
| Age (years) | Mean (SD) | 23.2 (5.98) |  | 24.3 (7.18) |  |
|  | Median | 21 |  | 22 |  |
|  | Min, Max | 18, 37 |  | 18, 40 |  |
| BMI | Mean (SD) | 19.8 (1.92) |  | 20.4 (1.67) |  |
|  | Median | 20.1 |  | 19.6 |  |
|  | Min, Max | 16.2, 23.1 |  | 18.8, 23.3 |  |
| Adolescents |  | n=33 |  | n=33 |  |
|  |  | N | % | N | % |
| Sex | Male | 17 | 52 | 15 | 45 |
|  | Female | 16 | 48 | 18 | 55 |
| PKDL grade | Grade 1 | 14 | 42 | 18 | 55 |
|  | Grade 2 | 15 | 45 | 12 | 36 |
|  | Grade 3 | 4 | 12 | 3 | 9 |
| Age (years) | Mean (SD) | 13.4 (1.78) |  | 13.0 (1.49) |  |
|  | Median | 12 |  | 12 |  |
|  | Min, Max | 12, 17 |  | 12, 17 |  |
| BMI | Mean (SD) | 17.2 (2.85) |  | 16.2 (2.01) |  |
|  | Median | 16.9 |  | 15.9 |  |
|  | Min, Max | 12.4, 26.9 |  | 12.9, 20.7 |  |

### Safety outcomes

There were 70 (25 local and 45 systemic) AEs reported by 43 participants during the study split 1.6:1 between vaccine and placebo groups (**Tables S2-S5)**. Average number of local AEs per participant were 0.30 in the vaccine vs 0.29 in the placebo arm (median of 0 in both arms), and systemic AEs per participant were 0.68 in the vaccine vs 0.36 in the placebo arm (median of 0 in both arms). Event numbers did not significantly differ between arms (Mann-Whitney U p =0.921 and p=0.501 respectively). AEs were limited to grade 1 and 2, with no grade 3, SAEs or SUSARs reported. All local and systemic AEs were deemed to be not serious and recovered. 44 AEs (25 local and 19 systemic) were considered possibly-, probably-, or definitely related to vaccination again showing a bias towards vaccine recipients (**Figure 2**). These included itch, pain or soft swelling at the injection site, headache, vomiting, fever and general muscle pain. Medication was not required for any of the local AEs but paracetamol (for headache, chills, fever) and chlorphenamine (for whole body itch) were prescribed for systemic AEs. No clinically relevant changes in blood biochemistry or haematology were observed. The most common systemic AE unrelated to the study was malaria (16 clinical episodes, 15 patients). One case of thrombocytopenia was recorded but deemed unrelated to vaccination (occurring in the placebo group).

**Figure 2.**
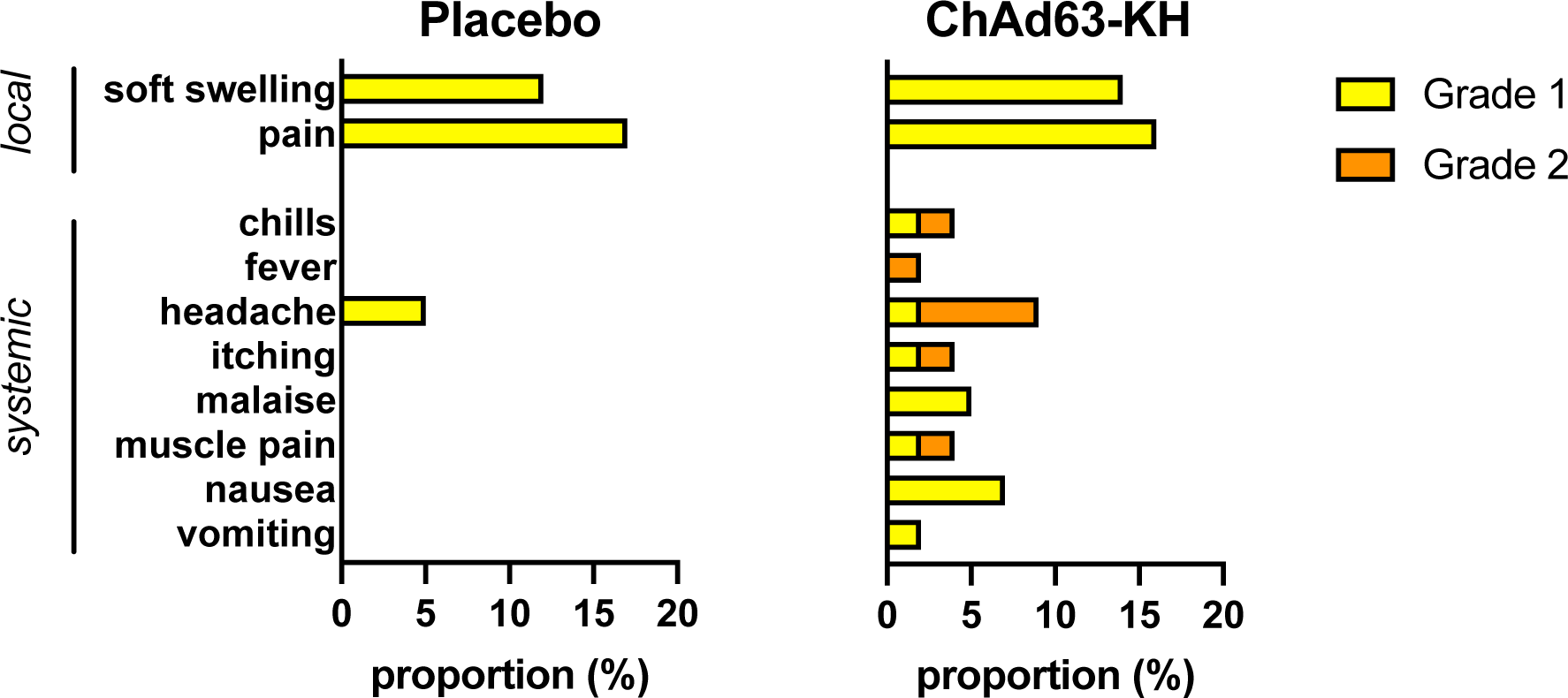
Adverse events associated with the LEISH2b trial. Data are shown for local and systemic adverse events judged to be possibly, probably, likely, or definitely associated with vaccination. Data are shown as proportion of participants in the study (n=86). Only grade 1 AEs (mild, yellow; 26 vs. 17 events overall in vaccine vs placebo group) and grade 2 AEs (moderate; orange; 17 vs. 10 events overall in vaccine vs. placebo group) were observed. See Tables S2-S5 for full details.

### Clinical outcomes

The per protocol primary outcome measure was 90% or greater improvement in clinical PKDL as determined by two clinical assessors. Of the 75 participants that completed the study to d90 (39F, 36M; **Table S1**), 6/40 (15%; 2F, 4M) and 4/35 (11%; 3F, 1M) in the vaccine and placebo arms respectively reached this threshold (RR 1.31 [95% CI, 0.40 to 4.28], p=0.742). To explore whether there was any vaccine effect at lower levels of improvement, we calculated the proportion of participants that attained 25, 50, 75 and 90% of improvement over time of follow up, and the trajectory of recovery appeared very similar between arms (**Figure 3a)**. Of note, all participants that reached the primary outcome did so between day 42 and d90 of follow up. The secondary outcome of PKDL grading (Grade 1 to Grade 4) was evaluated as PKDL severity is often used to monitor disease status ^33^. Grade distributions improved only marginally over time and were similar between arms (p =0.36, p= 0.53 and p = 0.38 for days 21, 42 and 90 respectively; Fisher’s exact test; **Figure 3b**).

**Figure 3.**
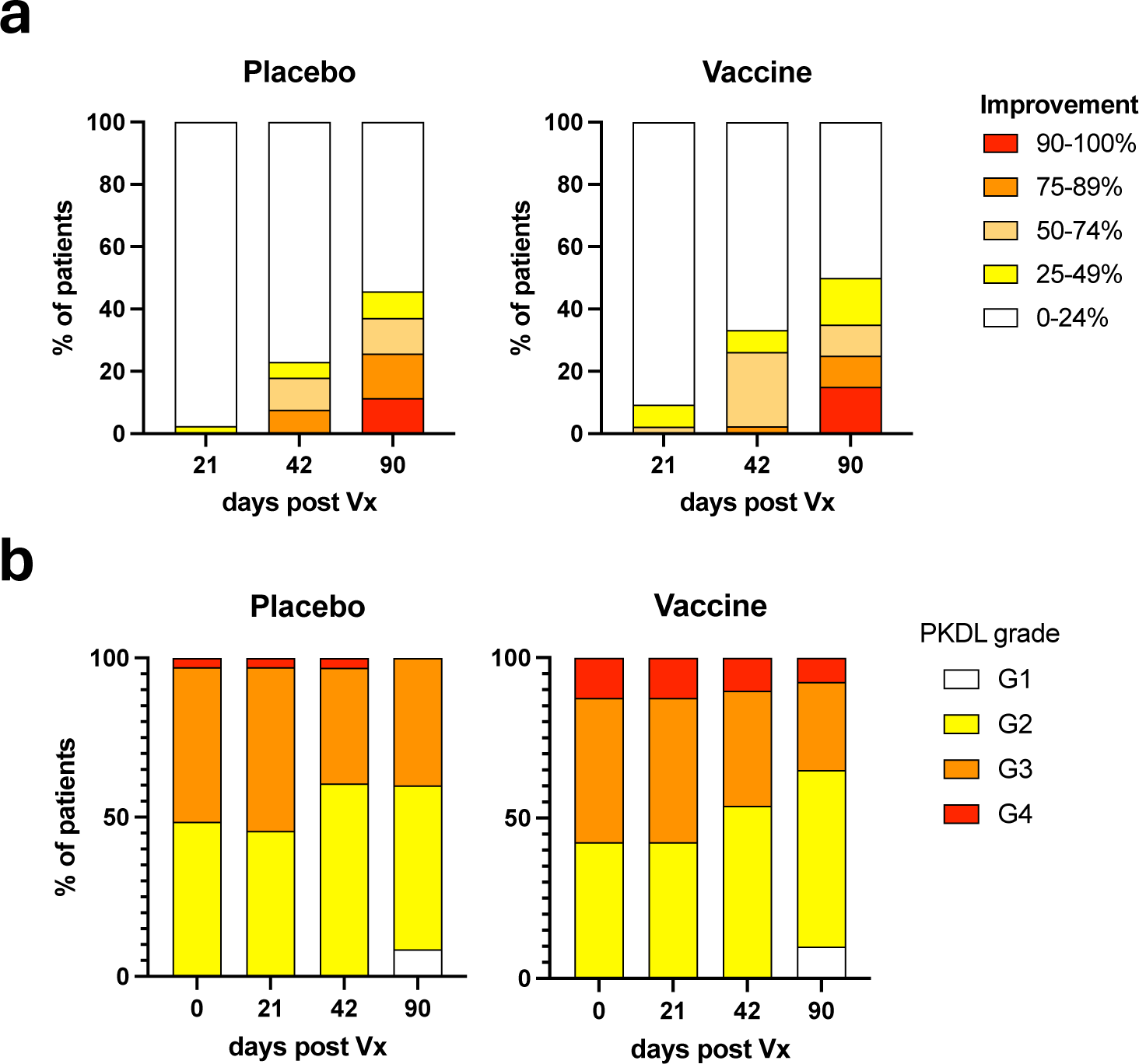
Clinical improvement in LEISH2b participants. **a.** Clinical improvement by study arm (n = 40 and n = 35 for vaccine and placebo groups respectively). Participants were categorised based on degree of improvement at different times post vaccination (Vx). Colour key indicates improvement category. **b.** Distribution of overall PKDL grades by study arm (n = 40 and n = 35 for vaccine and placebo groups respectively). Participants were scored for PKDL grade at vaccination (day 0) and at indicated days post vaccination (Vx). Colour key indicates grades. No significant differences were detected at any time point or over time (Fisher’s Exact test; see text).

As duration of PKDL has been suggested to influence cure rate ^19^, we additionally obtained prior duration in months data for a sub-group of 57/75 participants completing day 90 FU (45 adolescents, 12 adults; 33F, 24M). We stratified this subgroup into two classes based on duration of PKDL (<18 mths: median 11 months, range 6-11 months; n = 26 and ≥18mths, median 36 months, range 18 - 120 months, n= 31) and evaluated whether there was any difference in the proportion of patients reaching a conservative 50% improvement. In the <18 mths duration class, 6/11 receiving vaccine and 6/15 receiving placebo reached this threshold compared to 5/19 and 5/12 in the >18 mths duration class. Hence, duration of PKDL in this limited cohort did not appear to be associated with vaccine response or overall improvement.

Collectively, these analyses indicate that under the trial conditions employed, ChAd63-KH lacked efficacy as a single treatment in patients with persistent PKDL in Sudan.

### Whole blood transcriptome prior to and after vaccination

We used whole blood transcriptional analysis (WBTA) to confirm vaccine reactogenicity and capacity to induce immune responses in a subset of patients (n= 23 placebo and n= 27 Vx) for which PAXgene-collected blood samples were available for analysis. Compared to pre-vaccination, no differentially expressed genes (DEGs) were identified at day 1 post vaccination in patients receiving placebo (log2FC >1, adj p<0.05). In contrast in patients receiving ChAd63-KH, we identified 318 DEGs using this threshold (311 UP, 7 DOWN; **Table S6**). Using g-profiler, we annotated enriched terms associated with the response to ChAd63-KH, providing clear evidence of anti-viral, inflammatory, and immune response gene activation (**Figure 4a** and **Table S6**). To provide comparison with data from our previous Phase IIa study and responses to a range of other vaccines ^36^, we next identified transcriptional modules associated with vaccination ^37^. In keeping with the above analysis and with our previous analysis of ChAd63-KH responsiveness in PKDL patients ^31^, highly upregulated transcriptional modules included those associated with interferon responsiveness, dendritic cell and monocyte activation and antigen presentation (**Figure 4b** and **Table S6**). Out of 57 significantly enriched transcriptional modules, 31 were also enriched in the adolescent cohort of the LEISH2a study ^31^. Given that only 2/50 of the patients for which transcriptomic data was available achieved the primary outcome of ≥90% clinical response, it was not possible to conduct a robust analysis to identify differentially expressed genes or modules associated with clinical cure.

**Figure 4.**
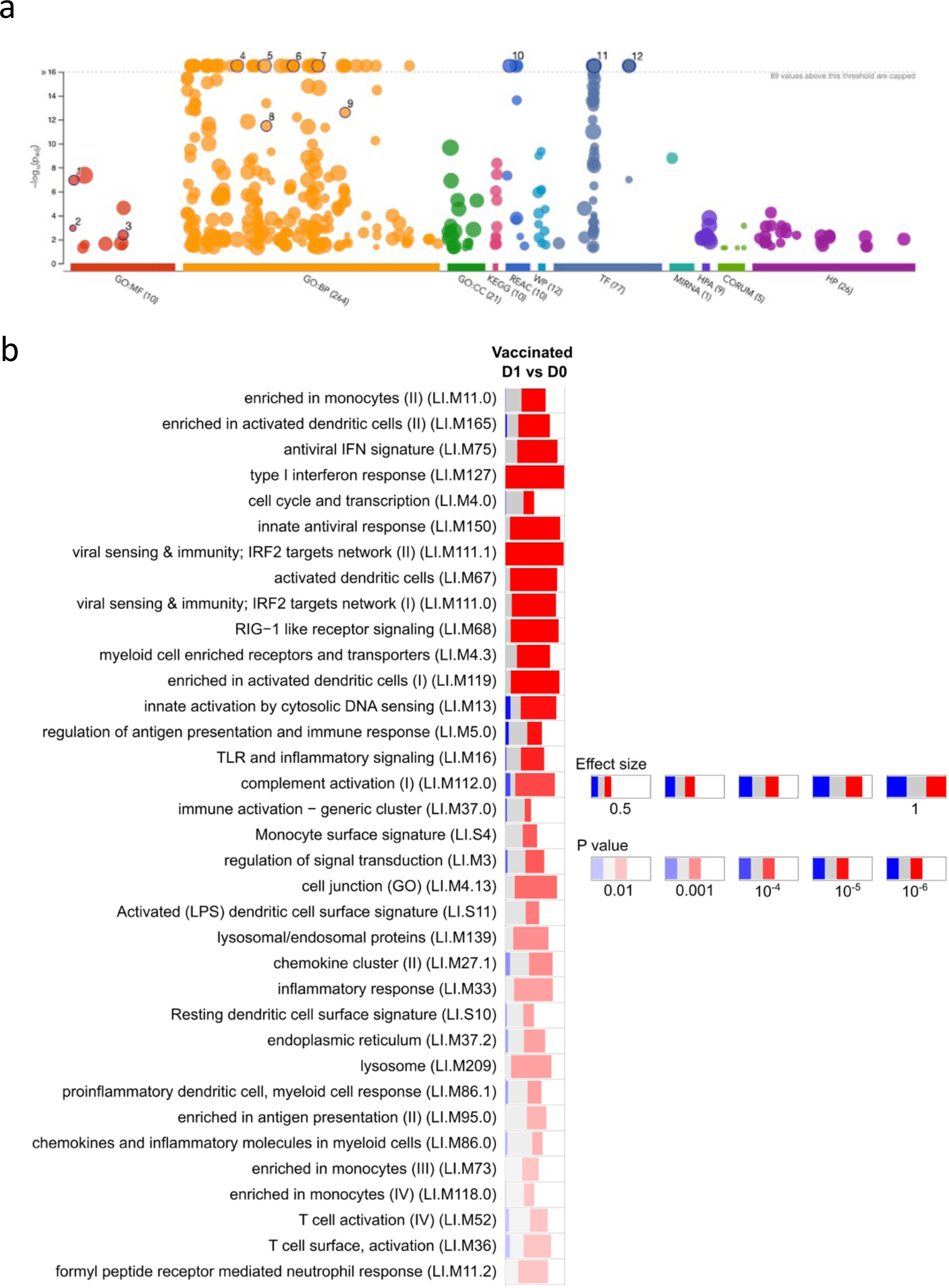
Whole blood transcriptomic analysis of vaccine response. WBTA was conducted on blood drawn prior to and 1 day post vaccination. Data are shown for 50 vaccinated participants (19F, 31M; 9 adults, 41 adolescents). **a**. gProfiler analysis of 311 upregulated genes. Representative enriched pathways are numbered: 1. GO:0003725, double stranded RNA binding; 2. GO:0001730, 2’-5’-oligoadenylate synthetase activity; 3. GO:0042379, chemokine receptor binding; 4. GO:0019221, cytokine-mediated signalling pathway; 5. GO:0034097, response to cytokine; 6. GO:0045089, positive regulation of innate immune response; 7. GO:0051607, defence response to virus; 8. GO:0034341, response to type II interferon; 9. GO:0071357, cellular response to type I interferon; 10. REAC:R-HAS-1, cytokine signalling in immune system; 11. TF:M00772, IRF motif,; 12. TF:M10080, STAT2 motif. Full details are provided in Table S6. **b.** Significantly enriched immune-related modules were identified applying the CERNO test on the adjusted p value-ranked lists of genes generated by DeSeq2 (see Table S6 for module gene lists). Bars represent proportion of significantly upregulated (red), downregulated (blue) or unchanged (grey) genes. The significance of module activation is proportional to the intensity of the bar, while the effect size is proportional to its width. Only top 35 modules (of 57; see Table S6) are shown for clarity. No DEGs and hence no modules were identified in patients receiving placebo.

## Discussion

ChAd63-KH is a “third generation” DNA vaccine based on adenoviral delivery of a gene construct encoding two well-characterised *Leishmania* antigens. In a Phase IIb stand-alone therapeutic trial in Sudanese patients with persistent PKDL, we have confirmed safety of this vaccine. However, this study failed to demonstrate clinical efficacy when measured by clearance of PKDL lesions over a 90-day follow up period.

The study was conducted in Gedaref state, South-eastern Sudan against a backdrop of significant adversity. Over the study period, Sudan sequentially experienced a revolution, a pandemic, a coup and military conflict that remains ongoing to date. Nevertheless, throughout this period, the trial team were able to maintain most aspects of trial management and governance although considerable delays in recruitment and in the ability to undertake sample analysis occurred. Following consultation with the DSMB and trial sponsor, the decision was taken in November 2022 to curtail recruitment at 86 participants, short of the 100 originally proposed. Full data analysis for the primary clinical outcome is reported here, with additional sub-group analysis being reported where patient metadata or biological samples were incomplete (either due to sample loss or data inaccessibility).

The primary clinical outcome of the study was to determine whether a single administration of ChAd63-KH vaccine was able to induce ≥90% improvement of PKDL in Sudanese patients with persistent disease of ≥ 6 months duration. Our analysis of 75 patients that that completed follow up to day 90 indicated that this was not the case. Similarly, PKDL grade was not improved over this time in vaccinated patients compared to those receiving placebo. Relaxing the criterion for clinical response also did not reveal any significant vaccine effect or effect of duration of prior PKDL. Hence, we conclude that this Phase IIb clinical trial has not demonstrated efficacy of ChAd63-KH under the conditions tested.

It is therefore important to consider the premise on which this study was based and potential reasons why efficacy was not observed. The immunopathogenic mechanisms that result in the chronic dermal presentation of PKDL are not fully understood. Immunohistological analysis of biopsies from Sudanese PKDL patients have noted the presence of CD4^+^ and CD8^+^ T cells but their functional state has not been characterised. Granuloma formation is seen in some cases but the histopathological picture may not correspond with clinical outcome ^23^. Keratinocyte IL-10 expression during VL was observed to be associated with later development of PKDL ^36^, but the specific role of IL-10 in PKDL per se is not understood. In Indian PKDL, more extensive studies have characterised CCR4^+^CD8^+^ T cells with a phenotype associated with exhaustion and immune regulation (PD-1^hi^ IL-10^+^) ^20^.

Collectively, these findings lend support to the hypothesis underpinning this trial i.e. that enhancing CD8^+^ T cell activation through vaccination may augment the ability of the host to clear parasites/antigens and reduce local pathology. Although we were unable to study antigen specific T cell responses in this study due to loss of all frozen cells, our previous studies have indicated that KMP-11- and HASPB-specific CD8^+^ IFNγ^+^ responses are induced in PKDL patients ^31^ and healthy volunteers ^30^ following ChAd63-KH vaccination. Given the comparability of the transcriptional response to vaccination seen in this and our other studies, we have no reason to suspect that similar T cell responses were not induced in this patient group. However, in the absence of efficacy, it is difficult to ascertain whether this CD8^+^ T cell response was merely insufficient in magnitude to elicit a clinical response or whether a broader response (in terms of antigen specificity or immune response quality) might be needed to achieve a clinical response.

With their known ability to rapidly induce antigen specific CD8^+^ T cells, adenoviral vectored vaccines have been extensively studied as potential therapeutic vaccines in cancer and chronic hepatitis B virus (HBV) infection. However, studies in these other diseases have to date also met with limited success. Whilst poor choice of immunogens was previously mooted as the explanation for therapeutic failure in cancer therapeutic trials, a common attribute of cancer and chronic viral disease is the development of a microenvironment that favours immune regulation or suppression. These changes often reflect the outcome of signalling through inhibitory checkpoint pathways (e.g. PD-1) or the induction of metabolic checkpoints such as IDO1, and it has been proposed that these disease-associated regulatory constraints on T cell function hinder vaccine efficacy. Experimental evidence to support this notion has been provided by Maini and colleagues who identified both NK cell and PD-1/PD-L1 signalling as negative regulators of HBV vaccine efficacy in animals ^37^. Whilst evidence for such an immunoregulatory environment is less well established in PKDL and other forms of dermal leishmaniasis, some similarities to these other chronic diseases exist. For example, both IDO1 and PD-L1 are abundant in chronic cutaneous leishmaniasis lesions resulting from *L. donovani* infection in Sri Lanka, with a reduction in PD-L1 expression predicting rate of cure after antimonial chemotherapy ^38^. More recently, we have identified expression of these checkpoint molecules in CL patients infected with *L.* (*V.*) *braziliensis* and also in PKDL patients infected with *L. donovani* in India ^39^. PKDL lesions are also enriched in M2 macrophages ^21^ contributing to a lack of leishmanicidal capacity and an immunoregulatory environment. Hence, it is possible that the local inhibitory environment in the skin of persistent PKDL patients provides an obstacle to effector CD8^+^ T cell function at this site. If this is indeed the case, it is likely that therapeutic vaccination in PKDL might also benefit from interventions that target regulatory pathways, such as those proposed in HBV ^37^. Whilst studies using existing biologics or other clinically approved drugs might provide proof of concept data in PKDL, it is recognised that such high-cost adjunct therapies are unlikely to drive vaccine implementation in a lower- and middle-income country (LMIC) setting. Alternatively, vaccination in conjunction with anti-leishmanial chemotherapy should also be considered, given previous successes with immunochemotherapy in PKDL ^29^ and more recent evidence suggesting that such chemotherapy may also diminish the immunoregulatory environment in CL patients^38^.

We confirm in this study the potent ability of ChAd63-KH to induce innate responses characterised by antiviral gene signatures and dendritic cell and monocyte activation, with weaker representation of modules associated with neutrophil response and complement activation. The modules signatures observed are also in keeping with those observed in a multi-vaccine analysis conducted by Hagan et al ^40^ which included an adenoviral vaccine candidate (MRKAd5-HIV) and with our previous analysis of PKDL patients ^31^. In the latter study, we observed that adolescents but not adults differentially expressed modules associated with B cell activation (M47.0 and M47.1). However, we did not observe differential expression these modules in the current study, despite the patient subgroup studied for WBTA being mainly composed of adolescents. The reasons for this difference are currently unknown and are subject to ongoing investigation. We had also previously identified 11 whole blood transcriptional modules predictive of 90% clinical cure, two of which were significantly differentially expressed between cure and non-cure participants (M139: lysosomal/endosomal proteins and M118.0: enriched in monocytes). Unfortunately, as only 2/50 of the samples analysed here achieved this level of clinical cure, we are unable to confirm or otherwise the predictive value of these modules.

In addition to the loss of samples / data due to the ongoing conflict in Sudan and other issues discussed above, a limitation in study design was the evaluation of only one vaccination schedule. ChAd63-KH was administered as a stand-alone therapy using a single dose given intramuscularly. This approach was chosen for both scientific and pragmatic reasons. Extensive data derived from experimental studies and early phase human trials ^41^ and from late phase trials and real world evidence obtained during the SARS-CoV-2 pandemic^42^ have indicated that repeat dosing with homologous adenovirus vaccines fails to significantly augment the CD8^+^ T cell response. Though direct evidence is lacking, it seems likely that a similar outcome would occur with vaccination in PKDL patients. Pragmatically, single dose schedules have multiple benefits in a LMIC setting, including reducing logistical complexities and a reduction in cost, not least due to the savings from not requiring manufacture of two clinical grade vaccines. Similarly, intramuscular dosing is preferred in many clinical situations due to ease of administration and standardisation and tolerability but may not be the most appropriate route for eliciting skin homing CD8^+^ T cells ^43^. Thus, we cannot rule out that an efficacy signal may have been observed using a repeat dosing schedule, by extending the period of follow up, by use of a heterologous prime-boost strategy ^41^ and / or by varying route of administration. Given the heterogeneity of clinical presentation and histopathology observed in PKDL, our data do not rule out the possibility of a therapeutic benefit from ChAd63-KH vaccination in either Sudanese patients with less persistent disease or in PKDL patients in South Asia. In addition, the prophylactic efficacy of this vaccine against different types of leishmaniasis remains to be evaluated. In conclusion, however, this Phase IIb study did not provide evidence to support progressing ChAd63-KH as a standalone therapeutic in Sudanese patients with persistent PKDL.

## Supporting information

Supplementary Data_Tables S2-S5 and SAP

## Data Availability

Processed gene expression data are available as supplementary material. Raw gene expression data will be available from GEO on publication.

## Contributors

Conceptualisation by EAGK, PMK, AML, CJNL, AMM and BMY. Clinical studies were conducted by BMY, EAGK, MO, AN, ETAE, A-AM, AJS, AMM, MA, ME, KS, and AEAM. Trial data collation and analysis was conducted by AK, RW, AMM, AML, CJNL, and PMK. Transcriptomic analysis was conducted by FS, CS, MN, GM, and PMK. Statistical analysis was conducted by AK and PMK. Writing original draft by PMK. Review & editing draft by CJNL, AML, AMM, BMY, RW and AK. Writing final draft by PMK. All authors read and approved the final version of the manuscript.

## Declaration of Interests

PMK and CJNL are co-inventors of a patent that covers the gene insert used in ChAd63-KH. No other conflicts of interest are declared.

## Acknowledgments

The clinical trial was funded by a Wellcome Trust Translation Award (WT108518MA; https://wellcome.ac.uk). Additional support for transcriptomics studies was provided by a Wellcome Trust Senior Investigator Award (to PMK; WT104726) and by the TRANSVAC2 program supported by the European Union’s Horizon 2020 Research and Innovation programme under grant agreement No. 730964 (TNA1802-02; https://www.transvac.org). GSK is allowing York University to use the cell line ProCell92 (proprietary of GSK) for their adeno program. GSK had the opportunity to review the publication, but the content is solely the responsibility of the authors. The authors also express their gratitude to the DSMB (Dr Rob Davidson, Dr Koert Ritmeijer, Dr Wolfgang Stohr, and Prof Alaadin Ahmed) for their advice, continual support and encouragement and to all the study participants and their families.

## Supplementary information

### Supplementary Data

The *Supplementary Data*_ *Tables S2-S5 and SAP file* contains the following:

1. Table S2 List of AEs by body system and preferred term
2. Table S3 List of AE severity by preferred term
3. Table S4 List of non-serious AEs
4. Table S5 List of vital signs and laboratory tests
5. Leish2b statistical analysis plan (SAP)

### Supplementary Data _Table S1

This *Excel file* supports Figure 3 and contains the following:

Raw data on PKDL improvement, prior PKDL duration, age, sex, subsequent treatment for all participants by coded ID.

### Supplementary Data_Table S6

This *Excel file* contains the following:

1. Raw counts table for 50 patients measured at day 0 and day 1 post vaccination
2. List of 311 upregulated genes in patients receiving ChAd63-KH (adj p<0.05; 2-fold change)
3. List of 7 down regulated genes in patients receiving ChAd63-KH (adj p<0.05; 2-fold change)
4. g-profiler output list of enriched terms for the 311 upregulated genes
5. Transcriptional module gene list (after Li et al (Ref 36))
6. Module analysis of response to ChAd63-KH

## Notes

### Clinical Trial

NCT03969134

### Author Declarations

The LEISH2b study (Clinicaltrials.gov ID: NCT03969134) was approved by the Sudan National Medicines and Poisons Board, and the Ethical Review Committees of the Institute of Endemic Diseases, University of Khartoum and the Department of Biology, University of York. LEISH2b was sponsored by the University of York. The study was conducted according to the principles of the current revision of the Declaration of Helsinki 2008 and ICH guidelines for GCP (CPMP/ICH/135/95). All participants provided written informed consent before enrolment. Consent forms are available accompanying the published protocol 32.

