## Supplementary Data_Tables S2-S5 and SAP for "A randomized double-blind Phase IIb trial to evaluate the efficacy of ChAd63-KH for the treatment of post kala-azar dermal leishmaniasis"

**Table S2: Incidence of Adverse Events by Body System and Preferred Term**

| Body System and Preferred Term | Group A | | | Group B | | |
|---|---|---|---|---|---|---|
| | Participants with any event n | Events n | Avg. events per participant (total: 44) | Participants with any event n | Events n | Avg. events per participant (total: 42) |
| **Overall** | **23** | **43** | **0.98** | **20** | **27** | **0.64** |
| **Local to injection site** | **13** | **13** | **0.30** | **12** | **12** | **0.29** |
| Pain/discomfort | 7 | 7 | 0.16 | 7 | 7 | 0.17 |
| Fluid/blood filled blisters | - | - | - | - | - | - |
| Soft Swelling | 6 | 6 | 0.14 | 5 | 5 | 0.12 |
| Hard Swelling | - | - | - | - | - | - |
| Redness/ discolouration | - | - | - | - | - | - |
| **Systemic** | **14** | **30** | **0.68** | **12** | **15** | **0.36** |
| Chills/rigor | 2 | 2 | 0.05 | - | - | - |
| Malaise/abnormal tiredness | 3 | 3 | 0.07 | - | - | - |
| General muscle ache | 2 | 2 | 0.05 | - | - | - |
| Fever | 1 | 1 | 0.02 | - | - | - |
| Headache | 6 | 6 | 0.14 | 3 | 3 | 0.07 |
| Nausea | 3 | 3 | 0.07 | - | - | - |
| Vomiting | 1 | 1 | 0.02 | - | - | - |
| Anaemia | 4 | 4 | 0.09 | - | - | - |
| Itching | 2 | 2 | 0.05 | - | - | - |
| Malaria | 6 | 6 | 0.14 | 9 | 10 | 0.24 |
| Thrombocytopenia | - | - | - | 1 | 1 | 0.02 |
| Tinea versicolor | - | - | - | 1 | 1 | 0.02 |

#### Table S3: Severity of Adverse Events by Preferred Term

| | Group A | | | Group B | | |
|---|---|---|---|---|---|---|
| **Body System and Preferred Term** | **Grade 1 n** | **Grade 2 n** | **Grade 3 n** | **Grade 1 n** | **Grade 2 n** | **Grade 3 n** |
| **Overall** | **26** | **17** | **0** | **17** | **10** | **0** |
| **Local to injection site** | **13** | **0** | **0** | **12** | **0** | **0** |
| Pain/discomfort | 7 | 0 | 0 | 7 | 0 | 0 |
| Fluid/blood filled blisters | - | - | - | - | - | - |
| Soft Swelling | 6 | 0 | 0 | 5 | 0 | 0 |
| Hard Swelling | - | - | - | - | - | - |
| Redness/ discolouration | - | - | - | - | - | - |
| **Systemic** | **13** | **17** | **0** | **5** | **10** | **0** |
| Chills/rigor | 1 | 1 | 0 | - | - | - |
| Malaise/abnormal tiredness | 2 | 1 | 0 | - | - | - |
| General muscle ache | 1 | 1 | 0 | - | - | - |
| Fever | 0 | 1 | 0 | 3 | 0 | 0 |
| Headache | 1 | 5 | 0 | - | - | - |
| Nausea | 3 | 0 | 0 | - | - | - |
| Vomiting | 1 | 0 | 0 | - | - | - |
| Anaemia | 0 | 4 | 0 | - | - | - |
| Itching | 1 | 1 | 0 | - | - | - |
| Malaria | 3 | 3 | 0 | 1 | 9 | 0 |
| Thrombocytopenia | - | - | - | 1 | 0 | 0 |
| Tinea versicolor | - | - | - | 0 | 1 | 0 |

#### Table S4: Non-Serious Adverse Events

*Sorted by group and days since vaccination*

| Participant ID | Gender | Days since vaccination | Description | Type | Relationship to Intervention | Grade | Outcome | Treatment |
|---|---|---|---|---|---|---|---|---|
| **Group A** | | | | | | | | |
| A002 | Female | 0 | Headache | Systemic | Probably related | Grade 2 | Recovered | Paracetamol |
| A002 | Female | 0 | Malaise | Systemic | Probably related | Grade 1 | Recovered | No |
| A002 | Female | 0 | Nausea | Systemic | Probably related | Grade 1 | Recovered | No |
| A002 | Female | 0 | Pain at injection | Local | Probably related | Grade 1 | Recovered | No |
| A004 | Male | 0 | Chills | Systemic | Probably related | Grade 1 | Recovered | No |
| A004 | Male | 0 | Pain at injection site | Local | Probably related | Grade 1 | Recovered | No |
| A006 | Female | 0 | General muscle pain | Systemic | Probably related | Grade 2 | Recovered | Paracetamol |
| A006 | Female | 0 | Nausea | Systemic | Probably related | Grade 1 | Recovered | No |
| A008 | Male | 0 | Soft swelling | Local | Probably related | Grade 1 | Recovered | No |
| A014 | Female | 0 | Soft swelling | Local | Probably related | Grade 1 | Recovered | No |
| A016 | Male | 0 | Pain at injection site | Local | Probably related | Grade 1 | Recovered | No |
| Y013 | Female | 0 | Soft swelling at injection site | Local | Probably related | Grade 1 | Recovered | No |
| Y021 | Male | 0 | Soft swelling | Local | Probably related | Grade 1 | Recovered | No |
| Y029 | Male | 0 | Nausea | Systemic | Probably related | Grade 1 | Recovered | No |
| Y031 | Female | 0 | Soft swelling | Local | Probably related | Grade 1 | Recovered | No |
| Y032 | Male | 0 | Pain at injection site | Local | Probably related | Grade 1 | Recovered | No |
| Y034 | Male | 0 | Pain at injection site | Local | Probably related | Grade 1 | Recovered | No |
| Y045 | Female | 0 | Soft swelling | Local | Probably related | Grade 1 | Recovered | No |
| Y050 | Male | 0 | Headache | Systemic | Possibly related | Grade 1 | Recovered | Paracetamol |

| Participant ID | Gender | Days since vaccination | Description | Type | Relationship to Intervention | Grade | Outcome | Treatment |
|---|---|---|---|---|---|---|---|---|
| A002 | Female | 1 | General muscle pain | Systemic | Probably related | Grade 1 | Recovered | No |
| A017 | Male | 1 | Pain at injection site | Local | Probably related | Grade 1 | Recovered | No |
| Y008 | Male | 1 | Headache | Systemic | Probably related | Grade 2 | Recovered | Paracetamol |
| Y008 | Male | 1 | Malaise | Systemic | Probably related | Grade 1 | Recovered | No |
| Y008 | Male | 1 | Pain at injection site | Local | Probably related | Grade 1 | Recovered | No |
| Y056 | Male | 1 | Chills/rigors | Systemic | Probably related | Grade 2 | Recovered | Paracetamol |
| Y056 | Male | 1 | Fever | Systemic | Probably related | Grade 2 | Recovered | Paracetamol |
| Y056 | Male | 1 | Headache | Systemic | Probably related | Grade 2 | Recovered | Paracetamol |
| Y056 | Male | 1 | Vomiting | Systemic | Probably related | Grade 1 | Recovered | No |
| A002 | Female | 3 | Itching all over the body | Systemic | Probably related | Grade 2 | Recovered | Chlorphenamine |
| A004 | Male | 10 | Itching all over the body | Systemic | Probably related | Grade 1 | Recovered | Chlorphenamine |
| Y029 | Male | 22 | Malaria | Systemic | Unrelated | Grade 2 | Recovered | Coartem |
| Y007 | Female | 96 | Malaria | Systemic | Unrelated | Grade 1 | Recovered | Coartem |
| Y063 | Male | 96 | Anaemia | Systemic | Unrelated | Grade 2 | Recovered | Ferrus Fumarate |
| Y049 | Female | 113 | Anaemia | Systemic | Unrelated | Grade 2 | Ongoing | Ferrous + Folic acid |
| Y049 | Female | 113 | Headache | Systemic | Unrelated | Grade 2 | Recovered | No |
| Y049 | Female | 113 | Malaise | Systemic | Unrelated | Grade 2 | Recovered | No |
| Y049 | Female | 113 | Malaria | Systemic | Unrelated | Grade 2 | Recovered | Coartem |
| Y004 | Female | 131 | Malaria | Systemic | Unrelated | Grade 1 | Recovered | Coartem |
| Y057 | Male | 133 | Anaemia | Systemic | Unrelated | Grade 2 | Ongoing | Folic Acid |
| Y053 | Male | 135 | Malaria | Systemic | Unrelated | Grade 1 | Recovered | Coartem |
| Y045 | Female | 139 | Anaemia | Systemic | Unrelated | Grade 2 | Ongoing | Ferrous + Folic acid |
| Y045 | Female | 139 | Headache | Systemic | Unrelated | Grade 2 | Recovered | No |
| Y045 | Female | 139 | Malaria | Systemic | Unrelated | Grade 2 | Recovered | Coartem |

| Participant ID | Gender | Days since vaccination | Description | Type | Relationship to Intervention | Grade | Outcome | Treatment |
|---|---|---|---|---|---|---|---|---|
| **Group B** | | | | | | | | |
| A003 | Male | 0 | Headache | Systemic | Probably related | Grade 1 | Recovered | No |
| A005 | Female | 40 | Headache | Systemic | Possibly related | Grade 1 | Recovered | Paracetamol |
| A005 | Female | 0 | Pain at injection site | Local | Probably related | Grade 1 | Recovered | No |
| A011 | Male | 23 | Malaria | Systemic | Unrelated | Grade 2 | Recovered | Coartem |
| A019 | Male | 132 | Malaria | Systemic | Unrelated | Grade 2 | Recovered | Coartem |
| A019 | Male | 0 | Pain at injection site | Local | Probably related | Grade 1 | Recovered | No |
| Y006 | Female | 96 | Malaria | Systemic | Unrelated | Grade 2 | Recovered | Coartem |
| Y010 | Male | 18 | Malaria | Systemic | Unlikely to be related | Grade 1 | Recovered | Coartem |
| Y015 | Female | 0 | Soft swelling at injection site | Local | Probably related | Grade 1 | Recovered | No |
| Y017 | Female | 0 | Soft swelling at injection site | Local | Probably related | Grade 1 | Recovered | No |
| Y022 | Male | 21 | Malaria | Systemic | Unrelated | Grade 2 | Recovered | Coartem |
| Y022 | Male | 96 | Malaria | Systemic | Unrelated | Grade 2 | Recovered | Coartem |
| Y022 | Male | 0 | Soft swelling | Local | Possibly related | Grade 1 | Recovered | No |
| Y023 | Female | 43 | Malaria | Systemic | Unrelated | Grade 2 | Recovered | Coartem |
| Y024 | Female | 0 | Soft swelling | Local | Probably related | Grade 1 | Recovered | No |
| Y027 | Male | 0 | Soft swelling | Local | Probably related | Grade 1 | Recovered | No |
| Y036 | Female | 0 | Pain at injection site | Local | Probably related | Grade 1 | Recovered | No |
| Y039 | Male | 0 | Pain at injection site | Local | Probably related | Grade 1 | Recovered | No |
| Y041 | Male | 139 | Malaria | Systemic | Unrelated | Grade 2 | Recovered | Coartem |
| Y042 | Male | 142 | Headache | Systemic | Unrelated | Grade 1 | Recovered | No |
| Y042 | Male | 142 | Malaria | Systemic | Unrelated | Grade 2 | Recovered | Coartem |
| Y042 | Male | 0 | Pain at injection site | Local | Probably related | Grade 1 | Recovered | No |
| Y047 | Male | 0 | Pain at injection site | Local | Probably related | Grade 1 | Recovered | No |

| Participant ID | Gender | Days since vaccination | Description | Type | Relationship to Intervention | Grade | Outcome | Treatment |
|---|---|---|---|---|---|---|---|---|
| Y058 | Male | 0 | Pain at injection | Local | Probably related | Grade 1 | Recovered | No |
| Y064 | Male | 96 | Tinea versicolor | Systemic | Unrelated | Grade 2 | Ongoing | Clotrimazole cream |
| Y065 | Female | 131 | Malaria | Systemic | Unrelated | Grade 2 | Recovered | Coartem Tabs |
| Y065 | Female | 131 | Thrombocytopenia | Systemic | Unrelated | Grade 1 | Ongoing | No |

#### Table S5: Vital Signs and Laboratory Tests Summary

**Raw Data**

| | | Group A | | | | | | Group B | | | | | |
|---|---|---|---|---|---|---|---|---|---|---|---|---|---|
| **Tests** | **Study Visits** | **N** | **Mean** | **SD** | **Min** | **Med** | **Max** | **N** | **Mean** | **SD** | **Min** | **Med** | **Max** |
| **Vital Signs** | | | | | | | | | | | | | |
| Blood Pressure Systolic | Screening | 44 | 106.55 | 7.053 | 100.0 | 110.0 | 120.0 | 42 | 105.95 | 6.270 | 90.0 | 110.0 | 120.0 |
| | Vaccination (before) | 44 | 108.41 | 8.337 | 100.0 | 110.0 | 130.0 | 42 | 107.98 | 8.700 | 90.0 | 110.0 | 120.0 |
| | Vaccination (2h after) | 44 | 108.18 | 8.700 | 90.0 | 110.0 | 120.0 | 42 | 106.31 | 6.989 | 100.0 | 105.0 | 120.0 |
| | Visit (D1) | 44 | 108.75 | 7.860 | 90.0 | 110.0 | 130.0 | 42 | 107.98 | 7.330 | 100.0 | 110.0 | 120.0 |
| | Visit (D3) | 44 | 108.86 | 7.840 | 90.0 | 110.0 | 120.0 | 42 | 107.98 | 8.268 | 90.0 | 110.0 | 120.0 |
| | Visit (D7) | 44 | 109.32 | 7.594 | 90.0 | 110.0 | 120.0 | 42 | 108.45 | 7.529 | 100.0 | 110.0 | 120.0 |
| | Visit (D21) | 43 | 107.21 | 7.966 | 90.0 | 110.0 | 130.0 | 41 | 107.32 | 7.340 | 100.0 | 110.0 | 120.0 |
| | Visit (D42) | 42 | 108.81 | 6.700 | 100.0 | 110.0 | 120.0 | 39 | 106.54 | 6.992 | 100.0 | 110.0 | 120.0 |
| | Visit (D90) | 40 | 108.75 | 9.111 | 100.0 | 110.0 | 140.0 | 35 | 106.71 | 7.568 | 90.0 | 110.0 | 120.0 |
| | Visit (D120) | 37 | 106.89 | 8.110 | 100.0 | 105.0 | 130.0 | 35 | 105.57 | 8.469 | 90.0 | 105.0 | 120.0 |
| Blood Pressure Diastolic | Screening | 44 | 67.73 | 7.270 | 60.0 | 70.0 | 80.0 | 42 | 67.02 | 6.055 | 60.0 | 70.0 | 80.0 |
| | Vaccination (before) | 44 | 70.11 | 7.662 | 55.0 | 70.0 | 90.0 | 42 | 70.00 | 6.533 | 60.0 | 70.0 | 80.0 |
| | Vaccination (2h after) | 44 | 68.64 | 6.413 | 60.0 | 70.0 | 80.0 | 42 | 68.10 | 7.321 | 55.0 | 70.0 | 80.0 |
| | Visit (D1) | 44 | 69.21 | 7.388 | 60.0 | 70.0 | 80.0 | 42 | 67.86 | 7.897 | 50.0 | 70.0 | 80.0 |
| | Visit (D3) | 44 | 68.43 | 7.825 | 55.0 | 70.0 | 80.0 | 42 | 68.69 | 6.631 | 60.0 | 70.0 | 80.0 |
| | Visit (D7) | 44 | 69.43 | 6.923 | 55.0 | 70.0 | 80.0 | 42 | 69.76 | 7.153 | 60.0 | 70.0 | 80.0 |
| | Visit (D21) | 43 | 67.67 | 7.665 | 60.0 | 70.0 | 80.0 | 41 | 69.15 | 7.323 | 60.0 | 70.0 | 80.0 |
| | Visit (D42) | 42 | 66.88 | 6.232 | 60.0 | 67.0 | 80.0 | 39 | 67.05 | 6.039 | 60.0 | 65.0 | 80.0 |
| | Visit (D90) | 40 | 67.75 | 7.067 | 60.0 | 67.5 | 90.0 | 35 | 65.71 | 5.443 | 60.0 | 65.0 | 80.0 |
| | Visit (D120) | 37 | 68.38 | 6.672 | 60.0 | 65.0 | 80.0 | 35 | 68.43 | 6.391 | 60.0 | 70.0 | 80.0 |
| Temperature | Screening | 44 | 36.46 | 0.362 | 35.8 | 36.4 | 37.1 | 42 | 36.44 | 0.454 | 35.6 | 36.4 | 38.1 |
| | Vaccination (before) | 44 | 36.33 | 0.565 | 35.0 | 36.4 | 37.3 | 42 | 36.36 | 0.478 | 35.4 | 36.4 | 37.4 |
| | Vaccination (2h after) | 44 | 36.51 | 0.389 | 35.2 | 36.5 | 37.1 | 42 | 36.51 | 0.380 | 35.6 | 36.5 | 37.1 |
| | Visit (D1) | 44 | 36.47 | 0.549 | 35.4 | 36.4 | 38.5 | 42 | 36.47 | 0.442 | 35.0 | 36.5 | 37.1 |
| | Visit (D3) | 44 | 36.52 | 0.513 | 35.4 | 36.6 | 37.3 | 42 | 36.51 | 0.395 | 35.5 | 36.5 | 37.3 |

| | | Group A | | | | | | Group B | | | | | |
|---|---|---|---|---|---|---|---|---|---|---|---|---|---|
| **Tests** | **Study Visits** | **N** | **Mean** | **SD** | **Min** | **Med** | **Max** | **N** | **Mean** | **SD** | **Min** | **Med** | **Max** |
| | Visit (D7) | 44 | 36.38 | 0.559 | 35.5 | 36.3 | 38.8 | 42 | 36.40 | 0.398 | 35.7 | 36.4 | 37.1 |
| | Visit (D21) | 43 | 36.46 | 0.365 | 35.7 | 36.5 | 37.2 | 41 | 36.45 | 0.447 | 35.7 | 36.4 | 37.4 |
| | Visit (D42) | 42 | 36.39 | 0.382 | 35.8 | 36.4 | 37.1 | 39 | 36.43 | 0.492 | 35.3 | 36.4 | 37.3 |
| | Visit (D90) | 40 | 36.57 | 0.417 | 35.8 | 36.6 | 37.7 | 35 | 36.51 | 0.395 | 35.7 | 36.5 | 37.3 |
| | Visit (D120) | 37 | 36.59 | 0.444 | 35.8 | 36.5 | 37.6 | 35 | 36.51 | 0.558 | 35.8 | 36.5 | 38.5 |
| Pulse | Screening | 44 | 80.11 | 6.229 | 68.0 | 80.0 | 100.0 | 42 | 81.79 | 5.629 | 68.0 | 81.0 | 92.0 |
| | Vaccination (before) | 44 | 76.71 | 6.025 | 65.0 | 76.0 | 100.0 | 42 | 79.10 | 7.077 | 70.0 | 77.0 | 98.0 |
| | Vaccination (2h after) | 44 | 78.50 | 6.245 | 60.0 | 79.0 | 90.0 | 42 | 79.86 | 7.118 | 60.0 | 80.0 | 98.0 |
| | Visit (D1) | 44 | 80.61 | 7.992 | 70.0 | 80.0 | 100.0 | 42 | 80.69 | 6.323 | 70.0 | 81.0 | 90.0 |
| | Visit (D3) | 44 | 79.30 | 5.564 | 70.0 | 80.0 | 92.0 | 42 | 80.21 | 6.107 | 70.0 | 80.0 | 92.0 |
| | Visit (D7) | 44 | 79.16 | 7.278 | 64.0 | 80.0 | 96.0 | 42 | 79.74 | 6.409 | 70.0 | 80.0 | 94.0 |
| | Visit (D21) | 43 | 80.09 | 5.433 | 68.0 | 80.0 | 90.0 | 41 | 78.95 | 6.008 | 60.0 | 80.0 | 88.0 |
| | Visit (D42) | 42 | 80.12 | 5.255 | 68.0 | 80.0 | 90.0 | 39 | 79.31 | 4.753 | 68.0 | 80.0 | 88.0 |
| | Visit (D90) | 40 | 80.63 | 5.977 | 68.0 | 80.0 | 90.0 | 35 | 80.80 | 5.764 | 68.0 | 80.0 | 90.0 |
| | Visit (D120) | 37 | 81.46 | 5.786 | 68.0 | 80.0 | 92.0 | 35 | 80.89 | 7.190 | 68.0 | 80.0 | 95.0 |
| Respiratory Rate | Screening | 44 | 17.86 | 2.216 | 14.0 | 18.0 | 24.0 | 42 | 18.05 | 1.899 | 14.0 | 18.0 | 22.0 |
| | Vaccination (before) | 44 | 18.98 | 2.029 | 16.0 | 18.0 | 24.0 | 42 | 18.76 | 2.184 | 14.0 | 18.0 | 24.0 |
| | Vaccination (2h after) | 44 | 18.91 | 2.301 | 12.0 | 20.0 | 22.0 | 42 | 18.64 | 1.885 | 14.0 | 18.0 | 24.0 |
| | Visit (D1) | 44 | 18.11 | 2.071 | 14.0 | 18.0 | 22.0 | 42 | 18.02 | 1.919 | 14.0 | 18.0 | 22.0 |
| | Visit (D3) | 44 | 18.36 | 2.616 | 12.0 | 18.0 | 25.0 | 42 | 18.00 | 2.306 | 14.0 | 18.0 | 22.0 |
| | Visit (D7) | 44 | 17.34 | 2.964 | 12.0 | 18.0 | 25.0 | 42 | 17.52 | 2.244 | 12.0 | 18.0 | 22.0 |
| | Visit (D21) | 43 | 17.74 | 2.205 | 12.0 | 18.0 | 24.0 | 41 | 18.02 | 2.219 | 12.0 | 18.0 | 22.0 |
| | Visit (D42) | 42 | 17.93 | 2.341 | 12.0 | 18.0 | 22.0 | 39 | 17.77 | 2.400 | 14.0 | 18.0 | 22.0 |
| | Visit (D90) | 40 | 17.28 | 2.025 | 14.0 | 18.0 | 22.0 | 35 | 17.60 | 2.291 | 14.0 | 18.0 | 22.0 |
| | Visit (D120) | 37 | 17.95 | 2.321 | 14.0 | 18.0 | 22.0 | 35 | 17.80 | 1.844 | 14.0 | 18.0 | 20.0 |
| **Biochemistry** | | | | | | | | | | | | | |
| ALT | Screening | 44 | 23.42 | 13.370 | 7.0 | 20.3 | 77.1 | 42 | 25.18 | 19.175 | 5.4 | 20.4 | 108.0 |
| | Visit (D1) | 44 | 23.51 | 14.573 | 6.6 | 20.4 | 78.0 | 42 | 23.49 | 16.299 | 11.6 | 19.7 | 93.0 |
| | Visit (D7) | 44 | 22.70 | 12.009 | 5.3 | 18.6 | 56.5 | 42 | 23.31 | 15.957 | 11.0 | 19.8 | 89.0 |
| | Visit (D90) | 40 | 24.00 | 12.692 | 7.0 | 20.1 | 66.3 | 35 | 21.40 | 9.321 | 9.2 | 19.2 | 53.0 |

| | | Group A | | | | | | Group B | | | | | |
|---|---|---|---|---|---|---|---|---|---|---|---|---|---|
| **Tests** | **Study Visits** | **N** | **Mean** | **SD** | **Min** | **Med** | **Max** | **N** | **Mean** | **SD** | **Min** | **Med** | **Max** |
| | Visit (D120) | 37 | 23.66 | 10.871 | 7.0 | 21.1 | 59.0 | 35 | 22.20 | 8.699 | 9.4 | 19.8 | 48.0 |
| AST | Screening | 44 | 27.88 | 10.989 | 10.5 | 26.8 | 60.7 | 42 | 27.06 | 13.486 | 11.4 | 23.7 | 71.0 |
| | Visit (D1) | 44 | 27.08 | 12.922 | 11.4 | 24.8 | 76.9 | 42 | 26.35 | 13.094 | 13.9 | 22.9 | 77.0 |
| | Visit (D7) | 44 | 25.39 | 8.016 | 12.5 | 23.2 | 49.4 | 42 | 25.68 | 11.925 | 14.0 | 23.8 | 78.0 |
| | Visit (D90) | 40 | 27.72 | 10.516 | 12.0 | 24.1 | 52.3 | 35 | 23.92 | 6.233 | 13.0 | 21.4 | 43.0 |
| | Visit (D120) | 37 | 28.43 | 10.745 | 9.6 | 28.0 | 66.0 | 35 | 26.52 | 7.377 | 13.0 | 23.4 | 44.0 |
| Creatinine | Screening | 44 | 0.47 | 0.203 | 0.10 | 0.41 | 1.10 | 42 | 0.42 | 0.170 | 0.19 | 0.40 | 0.90 |
| | Visit (D1) | 44 | 0.41 | 0.191 | 0.20 | 0.39 | 1.05 | 42 | 0.37 | 0.143 | 0.11 | 0.34 | 0.75 |
| | Visit (D7) | 44 | 0.39 | 0.142 | 0.20 | 0.35 | 0.80 | 42 | 0.37 | 0.176 | 0.14 | 0.35 | 0.94 |
| | Visit (D90) | 40 | 0.44 | 0.217 | 0.12 | 0.40 | 1.20 | 35 | 0.39 | 0.136 | 0.16 | 0.35 | 0.72 |
| | Visit (D120) | 37 | 0.42 | 0.199 | 0.18 | 0.40 | 1.10 | 35 | 0.37 | 0.142 | 0.10 | 0.40 | 0.73 |
| Albumin | Screening | 44 | 4.31 | 0.353 | 3.50 | 4.30 | 4.90 | 42 | 4.30 | 0.409 | 3.60 | 4.20 | 5.65 |
| | Visit (D1) | 44 | 4.21 | 0.333 | 3.70 | 4.20 | 4.90 | 42 | 4.17 | 0.380 | 3.40 | 4.15 | 5.10 |
| | Visit (D7) | 44 | 4.17 | 0.315 | 3.40 | 4.20 | 5.00 | 42 | 4.11 | 0.304 | 3.60 | 4.05 | 4.80 |
| | Visit (D90) | 40 | 4.23 | 0.335 | 3.60 | 4.20 | 4.80 | 35 | 4.15 | 0.323 | 3.60 | 4.10 | 4.70 |
| | Visit (D120) | 37 | 4.34 | 0.530 | 3.10 | 4.30 | 5.40 | 35 | 4.19 | 0.445 | 3.50 | 4.20 | 5.00 |
| Total Bilirubin | Screening | 44 | 0.58 | 0.270 | 0.19 | 0.55 | 1.60 | 42 | 0.60 | 0.343 | 0.20 | 0.52 | 1.40 |
| | Visit (D1) | 44 | 0.53 | 0.269 | 0.20 | 0.50 | 1.70 | 42 | 0.50 | 0.277 | 0.20 | 0.40 | 1.28 |
| | Visit (D7) | 44 | 0.54 | 0.244 | 0.20 | 0.48 | 1.16 | 42 | 0.55 | 0.296 | 0.20 | 0.49 | 1.44 |
| | Visit (D90) | 40 | 0.46 | 0.159 | 0.17 | 0.44 | 0.83 | 35 | 0.46 | 0.214 | 0.16 | 0.40 | 0.95 |
| | Visit (D120) | 37 | 0.56 | 0.271 | 0.20 | 0.52 | 1.30 | 35 | 0.51 | 0.291 | 0.12 | 0.44 | 1.20 |
| Direct Bilirubin | Screening | 44 | 0.13 | 0.117 | 0.00 | 0.10 | 0.60 | 42 | 0.17 | 0.212 | 0.01 | 0.10 | 0.96 |
| | Visit (D1) | 44 | 0.13 | 0.132 | 0.00 | 0.10 | 0.64 | 42 | 0.11 | 0.131 | 0.00 | 0.07 | 0.47 |
| | Visit (D7) | 44 | 0.10 | 0.082 | 0.00 | 0.09 | 0.30 | 42 | 0.11 | 0.104 | 0.00 | 0.09 | 0.50 |
| | Visit (D90) | 40 | 0.09 | 0.061 | 0.00 | 0.09 | 0.20 | 35 | 0.11 | 0.063 | 0.00 | 0.10 | 0.27 |
| | Visit (D120) | 37 | 0.15 | 0.177 | 0.00 | 0.10 | 0.88 | 35 | 0.12 | 0.124 | 0.00 | 0.10 | 0.60 |
| **Haematology** | | | | | | | | | | | | | |
| Haemoglobin | Screening | 44 | 13.32 | 1.233 | 9.9 | 13.2 | 16.8 | 42 | 13.11 | 0.877 | 11.4 | 13.1 | 15.6 |
| | Visit (D1) | 44 | 13.08 | 1.190 | 10.0 | 12.9 | 16.3 | 42 | 12.99 | 0.896 | 10.4 | 13.1 | 15.5 |
| | Visit (D7) | 44 | 13.22 | 1.227 | 9.9 | 13.1 | 16.5 | 42 | 13.08 | 0.989 | 11.2 | 13.0 | 16.2 |

| | | Group A | | | | | | Group B | | | | | |
|---|---|---|---|---|---|---|---|---|---|---|---|---|---|
| **Tests** | **Study Visits** | **N** | **Mean** | **SD** | **Min** | **Med** | **Max** | **N** | **Mean** | **SD** | **Min** | **Med** | **Max** |
| | Visit (D90) | 40 | 12.96 | 1.504 | 9.3 | 12.9 | 16.8 | 35 | 12.81 | 0.997 | 10.4 | 13.0 | 14.7 |
| | Visit (D120) | 37 | 12.60 | 1.673 | 8.5 | 12.4 | 16.2 | 35 | 12.70 | 1.154 | 9.5 | 13.0 | 14.5 |
| White blood cells | Screening | 44 | 6.02 | 1.621 | 3.6 | 5.9 | 10.2 | 42 | 6.49 | 1.544 | 3.3 | 6.5 | 11.5 |
| | Visit (D1) | 44 | 5.80 | 1.151 | 3.8 | 5.7 | 8.8 | 42 | 6.04 | 1.257 | 3.3 | 6.1 | 9.2 |
| | Visit (D7) | 44 | 5.70 | 1.217 | 3.3 | 5.6 | 9.7 | 42 | 6.03 | 1.517 | 3.4 | 5.8 | 10.5 |
| | Visit (D90) | 40 | 6.21 | 1.597 | 3.9 | 5.9 | 11.7 | 35 | 6.72 | 1.645 | 3.6 | 6.7 | 10.6 |
| | Visit (D120) | 37 | 5.86 | 1.416 | 4.1 | 5.6 | 9.6 | 35 | 6.00 | 1.578 | 3.8 | 5.8 | 10.0 |
| Neutrophils | Screening | 44 | 2.62 | 1.044 | 1.3 | 2.4 | 6.5 | 42 | 2.85 | 0.988 | 0.9 | 2.8 | 5.3 |
| | Visit (D1) | 44 | 3.22 | 1.039 | 1.5 | 3.1 | 5.9 | 42 | 2.77 | 0.933 | 1.4 | 2.7 | 5.2 |
| | Visit (D7) | 44 | 2.49 | 0.866 | 1.4 | 2.4 | 5.2 | 42 | 2.86 | 1.163 | 1.2 | 2.8 | 7.5 |
| | Visit (D90) | 40 | 2.79 | 1.429 | 1.4 | 2.5 | 10.0 | 35 | 3.26 | 1.053 | 1.2 | 3.4 | 6.7 |
| | Visit (D120) | 37 | 2.72 | 0.843 | 1.4 | 2.7 | 4.8 | 35 | 2.95 | 1.115 | 1.3 | 2.8 | 6.8 |
| Lymphocytes | Screening | 44 | 2.90 | 1.101 | 1.5 | 2.6 | 7.3 | 42 | 2.97 | 0.800 | 1.4 | 2.8 | 5.4 |
| | Visit (D1) | 44 | 2.01 | 0.625 | 0.8 | 1.9 | 3.5 | 42 | 2.66 | 0.595 | 1.3 | 2.6 | 3.8 |
| | Visit (D7) | 44 | 2.62 | 0.779 | 1.1 | 2.4 | 4.7 | 42 | 2.51 | 0.589 | 1.4 | 2.5 | 4.1 |
| | Visit (D90) | 40 | 2.74 | 0.855 | 1.3 | 2.8 | 5.2 | 35 | 2.71 | 0.901 | 0.2 | 2.6 | 5.0 |
| | Visit (D120) | 37 | 2.62 | 0.869 | 1.0 | 2.4 | 5.0 | 35 | 2.52 | 0.750 | 1.3 | 2.4 | 4.2 |
| Platelets | Screening | 44 | 296.59 | 66.361 | 178.0 | 296.0 | 437.0 | 42 | 332.24 | 99.642 | 158.0 | 327.0 | 638.0 |
| | Visit (D1) | 44 | 264.09 | 68.155 | 136.0 | 260.0 | 414.0 | 42 | 322.67 | 87.629 | 163.0 | 309.5 | 594.0 |
| | Visit (D7) | 44 | 280.14 | 66.329 | 153.0 | 292.0 | 426.0 | 42 | 323.64 | 85.216 | 143.0 | 303.0 | 491.0 |
| | Visit (D90) | 40 | 294.68 | 65.658 | 178.0 | 293.5 | 442.0 | 35 | 347.63 | 97.370 | 139.0 | 329.0 | 626.0 |
| | Visit (D120) | 37 | 290.14 | 77.797 | 176.0 | 273.0 | 470.0 | 35 | 320.23 | 99.209 | 86.0 | 309.0 | 526.0 |

**Change Since Screening**

| | | Group A | | | | | | Group B | | | | | |
|---|---|---|---|---|---|---|---|---|---|---|---|---|---|
| **Tests** | **Study Visits** | **N** | **Mean** | **SD** | **Min** | **Med** | **Max** | **N** | **Mean** | **SD** | **Min** | **Med** | **Max** |
| **Vital Signs** | | | | | | | | | | | | | |
| Blood Pressure Systolic | Screening | | | | | | | | | | | | |
| | Vaccination (before) | 44 | 1.86 | 6.561 | -10.0 | 0.0 | 20.0 | 42 | 2.02 | 7.656 | -10.0 | 0.0 | 20.0 |
| | Vaccination (2h after) | 44 | 1.64 | 6.796 | -20.0 | 0.0 | 10.0 | 42 | 0.36 | 6.931 | -10.0 | 0.0 | 20.0 |
| | Visit (D1) | 44 | 2.21 | 6.604 | -10.0 | 0.0 | 20.0 | 42 | 2.02 | 6.055 | -10.0 | 0.0 | 10.0 |
| | Visit (D3) | 44 | 2.32 | 5.556 | -10.0 | 0.0 | 20.0 | 42 | 2.02 | 6.055 | -10.0 | 0.0 | 20.0 |
| | Visit (D7) | 44 | 2.77 | 5.339 | -10.0 | 0.0 | 10.0 | 42 | 2.50 | 5.972 | -10.0 | 0.0 | 10.0 |
| | Visit (D21) | 43 | 0.98 | 7.042 | -18.0 | 0.0 | 20.0 | 41 | 1.46 | 5.836 | -10.0 | 0.0 | 10.0 |
| | Visit (D42) | 42 | 2.43 | 6.081 | -10.0 | 0.0 | 10.0 | 39 | 0.64 | 5.640 | -10.0 | 0.0 | 10.0 |
| | Visit (D90) | 40 | 2.30 | 6.509 | -10.0 | 0.0 | 30.0 | 35 | 1.29 | 5.333 | -10.0 | 0.0 | 10.0 |
| | Visit (D120) | 37 | 0.46 | 6.834 | -10.0 | 0.0 | 20.0 | 35 | -0.14 | 6.472 | -20.0 | 0.0 | 10.0 |
| Blood Pressure Diastolic | Screening | | | | | | | | | | | | |
| | Vaccination (before) | 44 | 2.39 | 7.662 | -10.0 | 0.0 | 20.0 | 42 | 2.98 | 6.154 | -10.0 | 0.0 | 20.0 |
| | Vaccination (2h after) | 44 | 0.91 | 6.313 | -10.0 | 0.0 | 20.0 | 42 | 1.07 | 5.795 | -10.0 | 0.0 | 15.0 |
| | Visit (D1) | 44 | 1.48 | 7.966 | -10.0 | 0.0 | 20.0 | 42 | 0.83 | 5.729 | -10.0 | 0.0 | 10.0 |
| | Visit (D3) | 44 | 0.71 | 7.593 | -10.0 | 0.0 | 20.0 | 42 | 1.67 | 5.372 | -10.0 | 0.0 | 20.0 |
| | Visit (D7) | 44 | 1.71 | 7.544 | -10.0 | 0.0 | 20.0 | 42 | 2.74 | 5.968 | -10.0 | 0.0 | 15.0 |
| | Visit (D21) | 43 | 0.23 | 7.396 | -20.0 | 0.0 | 20.0 | 41 | 2.07 | 6.223 | -10.0 | 0.0 | 10.0 |
| | Visit (D42) | 42 | -0.50 | 6.754 | -20.0 | 0.0 | 20.0 | 39 | 0.13 | 4.658 | -10.0 | 0.0 | 10.0 |
| | Visit (D90) | 40 | 0.25 | 6.197 | -10.0 | 0.0 | 20.0 | 35 | -1.29 | 4.902 | -10.0 | 0.0 | 10.0 |
| | Visit (D120) | 37 | 0.54 | 7.244 | -20.0 | 0.0 | 20.0 | 35 | 1.43 | 4.300 | -10.0 | 0.0 | 10.0 |
| Temperature | Screening | | | | | | | | | | | | |
| | Vaccination (before) | 44 | -0.13 | 0.721 | -2.0 | 0.0 | 1.1 | 42 | -0.08 | 0.645 | -1.5 | -0.1 | 1.3 |
| | Vaccination (2h after) | 44 | 0.06 | 0.547 | -1.8 | 0.1 | 1.1 | 42 | 0.07 | 0.651 | -2.5 | 0.1 | 1.2 |
| | Visit (D1) | 44 | 0.01 | 0.617 | -1.4 | 0.1 | 1.9 | 42 | 0.03 | 0.639 | -1.8 | 0.1 | 0.9 |
| | Visit (D3) | 44 | 0.06 | 0.531 | -1.1 | 0.1 | 1.1 | 42 | 0.08 | 0.551 | -1.2 | 0.2 | 0.9 |
| | Visit (D7) | 44 | -0.08 | 0.531 | -1.2 | -0.1 | 1.7 | 42 | -0.04 | 0.592 | -2.1 | 0.0 | 1.0 |
| | Visit (D21) | 43 | 0.01 | 0.491 | -0.9 | 0.0 | 1.0 | 41 | 0.02 | 0.536 | -1.0 | -0.1 | 1.0 |

| | | Group A | | | | | | Group B | | | | | |
|---|---|---|---|---|---|---|---|---|---|---|---|---|---|
| **Tests** | **Study Visits** | **N** | **Mean** | **SD** | **Min** | **Med** | **Max** | **N** | **Mean** | **SD** | **Min** | **Med** | **Max** |
| | Visit (D42) | 42 | -0.08 | 0.545 | -1.3 | 0.0 | 1.0 | 39 | -0.01 | 0.661 | -1.8 | 0.0 | 1.3 |
| | Visit (D90) | 40 | 0.12 | 0.504 | -1.0 | 0.3 | 0.9 | 35 | 0.06 | 0.552 | -1.7 | 0.1 | 0.9 |
| | Visit (D120) | 37 | 0.16 | 0.508 | -0.9 | 0.2 | 1.3 | 35 | 0.05 | 0.615 | -1.1 | 0.0 | 1.9 |
| Pulse | Screening | | | | | | | | | | | | |
| | Vaccination (before) | 44 | -3.41 | 7.913 | -20.0 | -2.5 | 28.0 | 42 | -2.69 | 7.511 | -18.0 | -4.0 | 18.0 |
| | Vaccination (2h after) | 44 | -1.61 | 8.911 | -20.0 | 0.0 | 16.0 | 42 | -1.93 | 7.906 | -20.0 | 0.0 | 14.0 |
| | Visit (D1) | 44 | 0.50 | 9.515 | -20.0 | 0.0 | 22.0 | 42 | -1.10 | 7.180 | -18.0 | 0.5 | 10.0 |
| | Visit (D3) | 44 | -0.82 | 7.711 | -18.0 | 0.0 | 16.0 | 42 | -1.57 | 7.232 | -18.0 | 0.0 | 12.0 |
| | Visit (D7) | 44 | -0.96 | 8.926 | -20.0 | 0.0 | 16.0 | 42 | -2.05 | 6.262 | -14.0 | -2.0 | 10.0 |
| | Visit (D21) | 43 | -0.30 | 7.704 | -24.0 | 0.0 | 16.0 | 41 | -2.88 | 6.218 | -14.0 | -4.0 | 8.0 |
| | Visit (D42) | 42 | 0.48 | 6.545 | -14.0 | 0.0 | 16.0 | 39 | -2.62 | 4.832 | -13.0 | -2.0 | 8.0 |
| | Visit (D90) | 40 | 0.75 | 7.232 | -18.0 | 0.0 | 16.0 | 35 | -1.97 | 6.392 | -20.0 | -2.0 | 12.0 |
| | Visit (D120) | 37 | 0.57 | 6.890 | -20.0 | 0.0 | 20.0 | 35 | -1.77 | 6.722 | -18.0 | 0.0 | 10.0 |
| Respiratory Rate | Screening | | | | | | | | | | | | |
| | Vaccination (before) | 44 | 1.11 | 1.907 | -2.0 | 2.0 | 6.0 | 42 | 0.71 | 2.266 | -2.0 | 0.0 | 9.0 |
| | Vaccination (2h after) | 44 | 1.05 | 2.362 | -4.0 | 1.5 | 6.0 | 42 | 0.60 | 2.275 | -4.0 | 0.0 | 6.0 |
| | Visit (D1) | 44 | 0.25 | 2.686 | -8.0 | 0.0 | 6.0 | 42 | -0.02 | 2.414 | -6.0 | 0.0 | 4.0 |
| | Visit (D3) | 44 | 0.50 | 2.445 | -8.0 | 0.0 | 7.0 | 42 | -0.05 | 2.603 | -6.0 | 0.0 | 6.0 |
| | Visit (D7) | 44 | -0.52 | 3.038 | -10.0 | 0.0 | 7.0 | 42 | -0.52 | 2.422 | -6.0 | 0.0 | 6.0 |
| | Visit (D21) | 43 | -0.16 | 2.400 | -8.0 | 0.0 | 6.0 | 41 | -0.02 | 2.329 | -5.0 | 0.0 | 4.0 |
| | Visit (D42) | 42 | -0.02 | 2.884 | -6.0 | 0.0 | 4.0 | 39 | -0.44 | 2.303 | -4.0 | 0.0 | 4.0 |
| | Visit (D90) | 40 | -0.63 | 2.382 | -6.0 | 0.0 | 4.0 | 35 | -0.51 | 2.582 | -6.0 | 0.0 | 4.0 |
| | Visit (D120) | 37 | 0.05 | 2.758 | -6.0 | 0.0 | 4.0 | 35 | -0.26 | 2.077 | -6.0 | 0.0 | 4.0 |
| **Biochemistry** | | | | | | | | | | | | | |
| ALT | Screening | | | | | | | | | | | | |
| | Visit (D1) | 44 | 0.08 | 6.812 | -23.4 | -0.5 | 29.1 | 42 | -1.69 | 6.291 | -28.9 | -0.7 | 11.0 |
| | Visit (D7) | 44 | -0.73 | 6.215 | -20.6 | -1.0 | 15.9 | 42 | -1.87 | 8.162 | -28.7 | -1.9 | 24.6 |
| | Visit (D90) | 40 | 0.47 | 11.291 | -30.2 | 0.2 | 42.2 | 35 | -5.00 | 15.054 | -55.0 | -1.8 | 11.8 |
| | Visit (D120) | 37 | -0.34 | 11.872 | -41.1 | 0.0 | 39.0 | 35 | -4.29 | 15.885 | -60.0 | -0.9 | 17.2 |
| AST | Screening | | | | | | | | | | | | |

| | | Group A | | | | | | Group B | | | | | |
|---|---|---|---|---|---|---|---|---|---|---|---|---|---|
| **Tests** | **Study Visits** | **N** | **Mean** | **SD** | **Min** | **Med** | **Max** | **N** | **Mean** | **SD** | **Min** | **Med** | **Max** |
| | Visit (D1) | 44 | -0.80 | 9.688 | -41.4 | -1.2 | 20.6 | 42 | -0.71 | 7.352 | -30.7 | -0.1 | 15.1 |
| | Visit (D7) | 44 | -2.49 | 8.277 | -39.1 | -0.7 | 10.8 | 42 | -1.39 | 6.415 | -27.7 | -0.2 | 8.0 |
| | Visit (D90) | 40 | -0.34 | 10.567 | -45.4 | 0.8 | 22.6 | 35 | -4.24 | 11.310 | -33.1 | -1.5 | 13.6 |
| | Visit (D120) | 37 | 0.04 | 12.288 | -31.2 | 1.0 | 35.0 | 35 | -1.70 | 11.939 | -34.0 | 0.6 | 18.0 |
| Creatinine | Screening | | | | | | | | | | | | |
| | Visit (D1) | 44 | -0.06 | 0.098 | -0.23 | -0.05 | 0.14 | 42 | -0.05 | 0.118 | -0.30 | -0.06 | 0.30 |
| | Visit (D7) | 44 | -0.08 | 0.168 | -0.57 | -0.06 | 0.22 | 42 | -0.05 | 0.139 | -0.35 | -0.05 | 0.26 |
| | Visit (D90) | 40 | -0.03 | 0.111 | -0.24 | -0.04 | 0.24 | 35 | -0.02 | 0.112 | -0.37 | 0.00 | 0.20 |
| | Visit (D120) | 37 | -0.06 | 0.142 | -0.44 | -0.02 | 0.20 | 35 | -0.04 | 0.146 | -0.30 | -0.07 | 0.23 |
| Albumin | Screening | | | | | | | | | | | | |
| | Visit (D1) | 44 | -0.10 | 0.272 | -0.70 | -0.10 | 0.80 | 42 | -0.13 | 0.318 | -1.35 | -0.10 | 0.80 |
| | Visit (D7) | 44 | -0.14 | 0.384 | -0.80 | -0.10 | 0.70 | 42 | -0.19 | 0.331 | -1.25 | -0.15 | 0.40 |
| | Visit (D90) | 40 | -0.10 | 0.385 | -1.00 | -0.10 | 0.50 | 35 | -0.16 | 0.530 | -1.95 | 0.00 | 0.70 |
| | Visit (D120) | 37 | 0.05 | 0.554 | -1.40 | 0.10 | 0.80 | 35 | -0.10 | 0.729 | -1.75 | 0.10 | 0.90 |
| Total Bilirubin | Screening | | | | | | | | | | | | |
| | Visit (D1) | 44 | -0.05 | 0.216 | -0.70 | -0.04 | 0.55 | 42 | -0.10 | 0.199 | -0.90 | -0.04 | 0.20 |
| | Visit (D7) | 44 | -0.04 | 0.213 | -0.58 | -0.01 | 0.52 | 42 | -0.05 | 0.294 | -0.90 | -0.01 | 1.12 |
| | Visit (D90) | 40 | -0.11 | 0.187 | -0.77 | -0.11 | 0.25 | 35 | -0.10 | 0.276 | -0.90 | -0.07 | 0.55 |
| | Visit (D120) | 37 | -0.02 | 0.221 | -0.80 | 0.00 | 0.45 | 35 | -0.05 | 0.275 | -0.79 | 0.01 | 0.42 |
| Direct Bilirubin | Screening | | | | | | | | | | | | |
| | Visit (D1) | 44 | 0.00 | 0.148 | -0.40 | 0.00 | 0.57 | 42 | -0.06 | 0.213 | -0.78 | 0.00 | 0.28 |
| | Visit (D7) | 44 | -0.02 | 0.102 | -0.30 | 0.00 | 0.15 | 42 | -0.07 | 0.229 | -0.90 | 0.00 | 0.25 |
| | Visit (D90) | 40 | -0.03 | 0.133 | -0.45 | -0.02 | 0.17 | 35 | -0.06 | 0.223 | -0.91 | 0.00 | 0.16 |
| | Visit (D120) | 37 | 0.03 | 0.193 | -0.41 | 0.02 | 0.79 | 35 | -0.06 | 0.243 | -0.76 | 0.00 | 0.58 |
| **Haematology** | | | | | | | | | | | | | |
| Haemoglobin | Screening | | | | | | | | | | | | |
| | Visit (D1) | 44 | -0.24 | 0.548 | -1.3 | -0.3 | 0.7 | 42 | -0.13 | 0.403 | -1.0 | -0.1 | 0.7 |
| | Visit (D7) | 44 | -0.10 | 0.590 | -1.4 | 0.0 | 1.0 | 42 | -0.03 | 0.648 | -1.4 | -0.1 | 1.7 |
| | Visit (D90) | 40 | -0.32 | 0.768 | -3.2 | -0.4 | 1.6 | 35 | -0.23 | 0.880 | -2.8 | -0.1 | 1.3 |
| | Visit (D120) | 37 | -0.70 | 1.022 | -3.5 | -0.6 | 1.2 | 35 | -0.35 | 0.960 | -2.8 | -0.3 | 1.7 |

| | | Group A | | | | | | Group B | | | | | |
|---|---|---|---|---|---|---|---|---|---|---|---|---|---|
| **Tests** | **Study Visits** | **N** | **Mean** | **SD** | **Min** | **Med** | **Max** | **N** | **Mean** | **SD** | **Min** | **Med** | **Max** |
| White blood cells | Screening | | | | | | | | | | | | |
| | Visit (D1) | 44 | -0.22 | 1.096 | -2.4 | -0.1 | 1.9 | 42 | -0.45 | 1.157 | -3.8 | -0.3 | 1.4 |
| | Visit (D7) | 44 | -0.32 | 1.050 | -3.1 | -0.4 | 2.2 | 42 | -0.46 | 1.343 | -4.2 | -0.5 | 2.9 |
| | Visit (D90) | 40 | 0.13 | 1.971 | -4.6 | 0.1 | 7.5 | 35 | 0.09 | 1.781 | -3.6 | 0.2 | 4.6 |
| | Visit (D120) | 37 | -0.23 | 1.387 | -2.6 | -0.4 | 2.7 | 35 | -0.55 | 1.467 | -3.9 | -0.6 | 2.9 |
| Neutrophils | Screening | | | | | | | | | | | | |
| | Visit (D1) | 44 | 0.61 | 0.951 | -1.5 | 0.5 | 3.6 | 42 | -0.08 | 0.818 | -2.5 | 0.1 | 1.7 |
| | Visit (D7) | 44 | -0.13 | 0.902 | -3.1 | -0.1 | 2.3 | 42 | 0.01 | 1.057 | -2.3 | 0.0 | 2.8 |
| | Visit (D90) | 40 | 0.14 | 1.744 | -3.5 | 0.1 | 7.7 | 35 | 0.34 | 1.169 | -1.4 | 0.1 | 3.2 |
| | Visit (D120) | 37 | 0.09 | 1.036 | -1.9 | 0.1 | 2.1 | 35 | 0.10 | 1.083 | -2.2 | 0.1 | 2.8 |
| Lymphocytes | Screening | | | | | | | | | | | | |
| | Visit (D1) | 44 | -0.89 | 0.997 | -5.4 | -0.7 | 1.2 | 42 | -0.31 | 0.643 | -2.3 | -0.2 | 0.7 |
| | Visit (D7) | 44 | -0.28 | 0.871 | -4.2 | -0.1 | 1.4 | 42 | -0.46 | 0.695 | -2.9 | -0.5 | 0.9 |
| | Visit (D90) | 40 | -0.09 | 0.658 | -1.7 | 0.0 | 1.3 | 35 | -0.36 | 0.949 | -2.8 | -0.3 | 1.4 |
| | Visit (D120) | 37 | -0.34 | 1.040 | -5.0 | -0.3 | 1.6 | 35 | -0.54 | 0.692 | -2.2 | -0.5 | 1.1 |
| Platelets | Screening | | | | | | | | | | | | |
| | Visit (D1) | 44 | -32.5 | 29.607 | -81 | -34.5 | 87.0 | 42 | -9.6 | 45.649 | -192 | -11.5 | 112.0 |
| | Visit (D7) | 44 | -16.5 | 33.533 | -104 | -17.0 | 64.0 | 42 | -8.6 | 44.338 | -147 | -12.5 | 111.0 |
| | Visit (D90) | 40 | -6.0 | 43.646 | -127 | -2.0 | 74.0 | 35 | 11.4 | 82.137 | -346 | 22.0 | 210.0 |
| | Visit (D120) | 37 | -6.9 | 51.279 | -190 | -10.0 | 87.0 | 35 | -16.7 | 66.440 | -238 | -3.0 | 72.0 |

### LEISH2b

### A phase IIb safety study to assess the safety and immunogenicity of a new Leishmania vaccine candidate ChAd63-KH

STATISTICAL ANALYSIS PLAN

Version 1.1

Version date: February 2024

(includes post-hoc exploratory analyses)

Author(s): Ada Keding

### 1 Introduction

#### 1.1 Scope of this Document

This statistical analysis plan (SAP) describes all intended analyses for the LEISH2b study data collected in the study database maintained by ClinServ. Statistical analysis of most study outcomes will be led by the statistics team at York Trials Unit, however the analysis of immunology data will additionally require the expertise of the immunology team at the Department of Biology at York University and will be detailed separately as appropriate.

#### 1.2 Version History

| Version | Date | Description / Specific Changes |
|---|---|---|
| 0.1 | June 2021 | First Draft |
| 0.2 | August 2022 | General Updates<br>Addition of explanation for non-stratified randomisation<br>Addition of interim primary outcome reveal to DSMB |
| 0.3 | February 2023 | General Updates<br>Confirmation of inclusion of ineligible screened participants<br>Confirmation of analysis of immunology data<br>Allowing for exclusion of patients from treatment analysis if no treatment was available due to medication shortage<br>Inclusion of suggested table/figure headings |

#### 1.3 Related Documentation

| Name | Description | Location | Latest Version | Date |
|---|---|---|---|---|
| Protocol | Full description of study design, procedures and definitions | Statistical Master File | 0.92 | 27/06/2019 |

#### 1.4 Glossary and Definitions

| | |
|---|---|
| AE | Adverse Event |
| BMI | Body Mass Index = weight / $height^2$ |
| CRF | Case Report Form |
| CTIMP | Clinical Trials of an Investigational Medicinal Product |
| DSMB | Data Safety Monitoring Board |
| DMP | Data Management Plan |
| IMP | Investigational Medicinal Product |
| IQR | Interquartile range |
| PKDL | Post kala-azar dermal leishmaniasis |
| SAE | Serious Adverse Event |
| SAP | Statistical Analysis Plan |
| SD | Standard Deviation |
| SMF | Statistical Master File |
| SOP | Standard Operating Procedure |
| SUSAR | Serious and Unexpected Adverse Reaction |
| TMF | Trial Master File |
| TSC | Trial Steering Committee |
| TMG | Trial Management Group |
| YTU | York Trials Unit |

### 2 Study Summary

#### 2.1 Background

Full details of the background to the trial and its design are presented in the latest protocol which is located in the Trial Master File (current protocol version at the time of writing is version 1.2, dated 22 December 2020).

#### 2.2 Design

LEISH2b is a randomised double-blind placebo-controlled phase 2b trial to assess the safety and efficacy of the Leishmania vaccine ChAd63-KH in patients with persistent PKDL in Sudan. Volunteer participants were randomly assigned to receive placebo or ChAd63-KH 7.5 x1010vp. Doses were administered at a single time point to patients under hospital care, and participants were followed up for 120 days after dosing.

#### 2.3 Study Objectives

Primary Objectives

- To assess the efficacy of the Leishmania vaccine ChAd63-KH compared with placebo. Efficacy was measured by the proportion of subjects at 42 and 90 days who do not require standard PKDL treatment.

- To assess the safety of the Leishmania vaccine ChAd63-KH in patients with persistent PKDL. Safety was measured by assessing adverse event data collected through history, clinical examination, blood tests and if necessary, tissue specific parasitological confirmation.

Secondary Objectives

- To observe any clinical changes in the cutaneous PKDL over a 120-day period following vaccination.
- To compare the humoral and cellular immune responses generated by the candidate vaccine in patients with persistent PKDL, compared with placebo-treated controls.

#### 2.4 Interventions

Participants received one of two preparations: a single intramuscular dose of

- ChAd63 KH $7.5 \times 10^{10}$ vp (The IMP was stored below -60°C. Prior to administration the IMP was removed from the dry ice and allowed to defrost to room temperature. Within one hour of the IMP being removed from the freezer it was administered to the volunteer)

or

- Placebo (normal saline)

The dose volume was injected into the deltoid muscle of the upper arm using a 21-23 gauge needle long enough to reach deep into the muscle. The needle was inserted at an angle of approximately 90° to the skin. Volunteers receiving the vaccine were asked which arm they would like to be injected.

#### 2.5 Sample Size

A total sample of size of 100 (randomly assigned 1:1 to be vaccinated with ChAd63-KH or placebo) is sufficient to detect an increase of ≥25% of PKDRL recovered patients (a reduction in patients of this magnitude requiring chemotherapy was judged to have clinical utility), assuming 90% power, 5%

statistical significance, a spontaneous clearance rate of ≤2% and loss to follow-up of ≤5%. This number of participants is typical for early phase vaccine studies.

The sample size was reviewed by the DSMB during the trial in light of a greater observed spontaneous clearance rate than was anticipated. Given the uncertainty around interim estimates of these rates, the continuation of the trial to the original target sample size was recommended.

##### 2.6 Enrolment

Otherwise healthy volunteers aged 8 – 50 years (age range updated in protocol version 1.1) with uncomplicated PKDL of more than 6 months' duration were approached to participate in the trial at the Professor El-Hassan's Centre for Tropical Medicine, Dooka, Gedarif State, Sudan. See protocol for full eligibility criteria and consent procedures.

##### 2.7 Randomisation and Blinding

A computer generated (Stata 16) randomisation list was prepared by the trial statistician, allocating participants 1:1 to either the active or placebo injection, using randomly permuted blocks, stratified by age group (adult or adolescent). The order was communicated to pharmacy staff at the study site through pre-prepared, sealed envelopes with sequential participant numbers.

During the course of the trial, adults were found to be more difficult to recruit, and the initial aim of an equal number of adolescents and adults in the trial was not feasible. However, the number of available randomisation envelopes in the Sudan was limited for each age group. Therefore, once allocations for the adolescent group had been used up, allocations for the adult stratum were used in order of presentation irrespective of age group, thus creating a single, unstratified randomised sample.

The vaccine and placebo injections were prepared in blacked out syringes labelled only with the participant identification number. The participants and clinical investigators (who administered the IMP and conducted the study follow-up) were blinded as to which injection patients received. The trial statistician (who generated the randomisation list), trial coordinator (who prepared sealed envelopes according to the randomisation list), assistant pharmacist (who prepared the IMP) and study nurse (who delivered the IMP to the clinical investigators) were not blind to the treatment allocation.

##### 2.8 Follow-up and Data Collection Schedule

All potential volunteers had a screening visit, which took place between 1 to 28 days before the vaccination. Informed consent was undertaken at the screening visit before any screening procedures. A day 0 visit was scheduled for the volunteer to receive their vaccination. Volunteers were required to stay in hospital for the first 3 days post vaccination. During this time, observations were performed at 30 minutes and 2hrs, and then at 24 hrs and 3 days after vaccination. Further visits took place at 7, 21, 42, 90 and 120 days after vaccination. These were considered compliant with the protocol if they took place +/-3 days either side of the target date (D21 and D42) or +/- 10 days (D90) or +/- 15 days (D120).

Table 1: Leish2b Data Collection Schedule

| | Screening | Vaccination[d] | In patient monitoring | In patient monitoring | Outpatient Follow up | Outpatient Follow up | Outpatient Follow up | Outpatient D90 | Outpatient D120 |
|---|---|---|---|---|---|---|---|---|---|
| Visit / Observation Number | 1 | 2 | 3 | 4 | 5 | 6 | 7 | 8 | 9 |
| Timeline (Days) | - | 0* | 1 | 3 | 7 | 21 | 42 | 90 | 120 |
| Time Window | -28 to -4 days | - | +/-3 hours | +/- 1 days | +/- 1 days | +/- 3 days | +/- 3 days | +/- 10 days | +/- 15 days |
| Medical History/Adverse event reporting | X | X | X | X | X | X | X | X | X |
| General Examination, vital signs | X | X[a] | X[a] | X[a] | X[a] | X[a] | X[a] | X | X |
| Height and weight | X | | | | | | | X | X |
| Examination of administration site | | X[b] | X | X | X | | | | |
| PKDL examination, grading & recording | X | X | X | X | X | X | X | X | X |
| Skin biopsy | X[c] | | | | | | | X[e] | |
| Urinalysis | X | X | | | | | X | X | X |
| Urinary pregnancy test (females only) | X | X[d] | | | X | | X | | |
| Haematology & Biochemistry (5ml) | X | X[d] | X | | X | | | X | X |
| Blood Borne Virus Screen(2.5ml) | X | | | | | | X | | |
| Bio-Rad LEISH IT (Leishmania antibody test) (1ml) | X | | | | | | | | |
| Microarray bloods (2.5ml) | | X[f] | X | X | X | | | | |
| Cellular Responses (10ml) | | X | | | | X (15ml) | X | X | X |
| Serum (2.5ml) | | X | | | | | X | | |
| Malaria RDT (0.5ml) | X | | | | | X | X | X | X |
| Blood Volume Per Visit | 9.0ml | 20.0ml (15ml in 8-11yrs) | 7.5ml | 2.5ml | 7.5ml | 15.5ml | 15.5ml | 15.5ml | 15.5ml |

* admission to hospital day before vaccination day

a Full general examination only if required

b At vaccination, examination of administration site and recording of observations will be done at 10 minutes, 60 minutes and 120 minutes post vaccination.

c No skin biopsy at entry in 8-11 yr olds

d. Repeat haematology and biochemistry, as well as repeat urine pregnancy test if female, do not need to be performed if vaccination is within 48 hours of the initial or repeat screening tests, or in 8-11 yr olds.

e Inpatients receiving SOC treatment as an inpatient

f Microarray blood sampling will be performed pre-vaccination.

##### 2.9 Study Outcomes

###### 2.9.1 Primary Outcomes

- Efficacy: PKDL Clearance rate at day 42 and day 90
    - The severity, extent and distribution of the PKDL disease was measured by a standardized grading system and image capture
    - If there was a <75% improvement in the degree of PKDL then the patient was offered standard PKDL treatment of 20 days liposomal Amphotericin B. If there was a 75-90% improvement in the degree of PKDL then the CI in consultation with the patient decided on conservative treatment or Amphotericin B. With >90% improvement no further intervention was recommended.
    - Patients were considered PKDL recovered with ≥90% PKDL clearance, i.e. requiring no further treatment

- Safety: Median number of local (at the injection site) and systemic events per patient
    - Adverse events were collected throughout the follow-up period through medical history, direct questioning, clinical examination, blood tests and if necessary, tissue specific parasitological confirmation
    - Adverse events were graded in their severity and likely relationship with IMP according to the study protocol
    - For laboratory parameters, set ranges identified normal as well as mild, moderate, severe and extreme adverse event bands. Based on CI opinion, out of range parameters were classified as clinically significant AEs or non-clinically significant AEs.

###### 2.9.2 Secondary Outcomes

Clinical PKDL Changes

- PKDL Recovery with ≥90% PKDL clearance at day 120
- PKDL Recovery with ≥75% PKDL clearance at day 42, day 90 and day 120
- Agreed treatment with liposomal Amphotericin B at day 90
- Areas of the body involved (Face, Chest, Back, Arms, Legs) and grade by area at all time points (day 1, day 3, day 7, day 21, day 42, day 90, day 120)
    - Grade 0: no lesion
    - Grade 1 single lesion
    - Grade 2: multiple lesions
    - Grade 3: multiple coalesced lesions
- Body part total grade score at all time points (day 1, day 3, day 7, day 21, day 42, day 90, day 120), range 0 to 15
- Overall lesions grade
    - Grade 0: none
    - Grade 1: Scattered maculopapular or nodular lesions, covering mainly the face particularly around the mouth and eyes
    - Grade 2: Dense maculopapular or nodular rash covering most of the face and extending to chest, back, upper arms and legs
    - Grade 3: Dense maculopapular or nodular rash covering most of the body, including hands and feet

Immune Response

- Measures of T cell, B cell and innate immunity induced by the vaccine e.g. as measured by
    - ELISPOT
    - Flow cytometry
    - ELISA and/or transcriptomics

### 3 Data Management

##### 3.1 Case Report Form (CRF) Data

Data are collected in paper form at the study site.

###### 3.1.1 Data Sources

- Screening log
- Screening CRF
- Vaccination CRF
- Day 1, 7, 21, 42, 90, 120 CRFs
- Unscheduled Visit CRF
- Concomitant Medications CRF
- Adverse Event CRF

###### 3.1.2 Data Receipt and Entry

- CRFs will be photocopied and the originals sent to ClinServ for data management
- Data will be received, logged and entered into a study database
- Data are be checked, queried and cleaned by the ClinServ Data Manager
- Data are stored in a central, secure access database

###### 3.1.3 Data Transfer

- A copy of the study database will be made available to the Trial Statistician upon request in a spreadsheet format
- The Statistician will import the relevant datasets for analysis using the latest version of Stata and save this in the SMF
- A copy of the CRFs marked up with the variable names used in the final Stata data set will be kept by the trial statistician in the SMF

###### 3.1.4 Data Queries

- The Statistician will perform data checks (verification, consistency, range checks, missing data) and variables will be examined for unusual, outlying, unlabelled or inconsistent values.
- Queries identified by the statistician during the analysis will be sent to the trial team / the ClinServ Data Manager as appropriate
- Queries, responses and any necessary database updates will be logged

##### 3.2 Laboratory Data

Laboratory / immunology data will be sent securely and anonymously to the Data Manager. The expected file format is Excel at time of writing. The data will be analysed as directed by colleagues in the Biology department at the University of York. The analysis of the laboratory data is not included in the scope of this analysis plan.

### 4 Interim Analyses

There were no planned interim analyses, however the DSMB could request unblinded outcome data at any time.

In June 2021, concerns emerged regarding higher spontaneous cure rates than the anticipated 2% in the sample size, which could result in an underpowered analysis given the recruitment progress at the time. As a result, the primary outcome was presented to the DSMB for the placebo group only in October 2021 and possible scenarios for effect sizes and achieved power for different numbers of recruited participants outlined. It was decided that the presented cure rate was not a reliable estimate owing to the small sample size, and that under plausible recruitment and cure rate scenarios sufficient statistical power remained to recommend the continuation of the study.

### 5 End of Study Statistical Analysis

#### 5.1 Reporting Timelines

The Statistician will provide a final report after data have been collected and entered for all patients and all data queries have been resolved. The last time point for data collection will be when the last patient vaccinated has reached 120 days of follow up. It is anticipated that the Trial Statistician will produce a report within 6 months after this date, allowing for data entry, checking and validation.

#### 5.2 Report Format and Assumptions

All analyses will be performed using the latest version of Stata (Version 17) and the version number will be recorded in trial reports.

Descriptive results will be presented for all patients, by randomised group and further broken down by age group (adults or adolescents). Whilst comparisons between the two age groups may be detailed, there is no aim to demonstrate statistically significant differences from such a comparison.

All analyses will be carried out on available patient data, and no imputations for missing data will be conducted.

The study statistician will not be blind to treatment allocation, however the analysis will be checked by a further independent statistician before the release of results.

#### 5.3 Study Progress

A CONSORT diagram displaying the flow of participants through the trial will be completed. The number of screened patients and reasons for exclusion will be detailed. Reasons for missing data and non-completion of the study will be detailed by randomised group. Any protocol deviations will be reported.

*Figure 1: Recruitment over time*
*Figure 2: Patient flow through trial (CONSORT)*
*Table 1: Reasons for non-completion*

#### 5.4 Baseline Characteristics

All collected data (age, gender, BMI, general symptoms, vital signs, biochemistry etc.) at screening will be summarised descriptively for: all patients, by randomised group and by age group. Continuous measures will be reported as averages (n, mean, standard deviations, median, min, max) while categorical data will be reported as counts and percentages. No formal statistical comparisons will be undertaken to compare baseline data between groups.

*Table 2: Baseline Characteristics by Allocation*
*Table 3: Baseline Characteristics by Age Group*

#### 5.5 Follow-up Data

All collected volunteer data will be reported in a similar format to baseline characteristics with the addition of change over time by trial arm, age group and total number of volunteers for the vaccination visit and all follow-up visits at day 1, 3, 7, 21, 42, 90 and day 120. Missing data will be reported for each outcome.

*Table 4: Vital Signs, Biochemistry, Haematology by Allocation (Total)*
*Table 5: Change in Vital Signs, Biochemistry, Haematology by Allocation (Total)*
*Table 6: Vital Signs, Biochemistry, Haematology by Allocation (Adolescents)*
*Table 7: Change in Vital Signs, Biochemistry, Haematology by Allocation (Adolescents)*
*Table 8: Vital Signs, Biochemistry, Haematology by Allocation (Adults)*
*Table 9: Change in Vital Signs, Biochemistry, Haematology by Allocation (Adults)*

#### 5.6 Analysis of Primary Outcomes

##### 5.6.1 Efficacy Outcomes: PKDL recovery

PKDL clearance rates (≥75% and ≥90% improvement) will be presented descriptively at day 42, day 90 and at day 120 by trial arm (counts and percentages).

The primary outcome is the proportion of PKDL recovered patients at day 42 and day 90 with ≥90% improvement, i.e. not requiring treatment. In the primary analysis, logistic regressions will be used to analyse treatment differences between randomised trial arms (vaccine versus placebo) at day 42 and day 90, additionally adjusting for age group. Relative risk ratios (for the 'risk' of recovery) will be presented with 95% confidence intervals and p-values. If the recovery event rate is very low (fewer than 10 PKDL recovered patients), a more simple, unadjusted Chi Squared or Fisher's Exact test (depending on test assumptions) will be chosen for the analysis instead.

*Table 10: PKDL Recovery Rates over Time by Allocation*
*Figure 3: Bar chart of PKDL Recovery over Time by Allocation (≥90% improvement)*

##### 5.6.2 Safety Outcomes: Adverse Events

The number and proportion of adverse events, categorised by Body system (local or systemic), Preferred term, Seriousness, Grade and Relatedness to the study treatment will be presented by trial arm and age group, separately for serious and non-serious adverse events.

The median number of serious adverse events and non-serious adverse events per participant (separately for local and systemic events) will be compared between randomised groups using the Mann-Whitney-U test and the p-value presented. The distribution of number of adverse events per participant will be illustrated using box plots.

A full list of adverse events including event descriptions will be presented in order of time since randomisation.

*Table 10: Summary of Serious Adverse Events by Allocation (Total)*
*Table 11: Summary of Non-Serious Adverse Events by Allocation (Total)*
*Figure 4: Box Plot of Number of Serious Adverse Events per Patient by Allocation (Total)*
*Figure 5: Box Plot of Number of Non-serious Adverse Events per Patient by Allocation (Total)*
*Table 12: Summary of Serious Adverse Events by Allocation (Adolescents)*
*Table 13: Summary of Non-Serious Adverse Events by Allocation (Adolescents)*
*Table 14: Summary of Serious Adverse Events by Allocation (Adults)*
*Table 15: Summary of Non-Serious Adverse Events by Allocation (Adults)*
*Table 16: Comparison of Adverse Events per Patient by Allocation*

*Table 17: Full Listing of all Adverse Events*

#### 5.7 Analysis of Secondary Outcomes

##### 5.7.1 PKDL recovery

PKDL recovery at rates / times that did not constitute the primary outcome (≥90% clearance at day 120 and ≥75% PKDL clearance at day 42, day 90 and day 120) will be compared between vaccine and placebo as the primary outcome with logistic regressions, chi-square, or Fisher's exact test as appropriate.

*Table 10: PKDL Recovery Rates over Time by Allocation*
*Figure 3: Bar chart of PKDL Recovery over Time by Allocation (≥90% improvement)*
*Figure 4: Bar chart of PKDL Recovery over Time by Allocation (≥75% improvement)*

##### 5.7.2 Agreed Treatment

In addition to the recovery estimated by percentage PKDL clearance, the requirement for treatment will be assessed by differences in the proportion of agreed treatment with liposomal Amphotericin B at day 90. Patients for whom no treatment was available due to medication shortage will be excluded from this analysis. Treatment rates will be compared between vaccine and placebo as the primary outcome with logistic regressions, chi-square, or Fisher's exact test as appropriate.

*Table 10: PKDL Recovery Rates over Time by Allocation*

##### 5.7.3 Changes in PKDL appearance

Overall PKDL lesion grade (Grade 0 to 3) will be presented descriptively (counts and percentages) by trial arm at all available time points in total and by age group.

PKDL lesion grade (Grade 0 to 3) for each body part will be presented descriptively (counts and percentages) together with the overall sum grade score adding up all body parts (score 0 to 15, means and SDs) by trial arm at all available time points in total and by age group.

Only overall PKDL lesion grade will be compared statistically between groups using Fisher's Exact test.

*Table 18: PKDL Overall Lesions Grade over Time by Allocation (Total)*
*Table 19: PKDL Lesions Grade by Body Part over Time by Allocation (Total)*
*Table 20: PKDL Body Part Lesion Sum Score over Time by Allocation (Total)*
*Table 21: PKDL Overall Lesions Grade over Time by Allocation (Adolescents)*
*Table 22: PKDL Lesions Grade by Body Part over Time by Allocation (Adolescents)*
*Table 23: PKDL Body Part Lesion Sum Score over Time by Allocation (Adolescents)*
*Table 24: PKDL Overall Lesions Grade over Time by Allocation (Adults)*
*Table 25: PKDL Lesions Grade by Body Part over Time by Allocation (Adults)*
*Table 26: PKDL Body Part Lesion Sum Score over Time by Allocation (Adults)*

##### 5.7.4 Immune Response

KMP HASPB antibody responses will be assessed by serum ELISA and T cell responses to the KMP-HASP protein by ELISpot. Groups will be allocated by arm (vaccine versus placebo) and summary statistics from the ELISPOT and ELISA responses from baseline and post-vaccination until 120 days (day 0, 21, 42, 90 and 120) will be illustrated. Microarray bloods will be sent to the University of Siena for transcriptomic analysis.

*Planned output to be detailed separately outside of this document*

##### 5.8 Post-hoc Exploratory Analyses

Following completion of follow-up and the availability of study results, the following additional exploratory analyses were conducted in order to further quantify the magnitude and timing of recovery rates and explore reasons for lack of observed efficacy:

- PKDL improvement was categorised into more gradual thresholds than the 90% binary clearance rate (0-24%, 25-49%, 50-74%, 75-89%, 90-100%), and the recovery profile was compared between randomised groups at available time points

- It is possible that duration of PKDL may have influenced cure rate. Therefore baseline duration of PKDL was obtained for study participants where possible and a moderate 50% improvement rate was compared between randomised groups for shorter (less than median) and longer (greater than median) PKDL duration

### 6 Signatures of Approval

| Name | Trial Role | Signature | Date |
|---|---|---|---|
| Ahmed Musa | Chief Investigator<br>Institute of Endemic Diseases<br>University of Khartoum, Sudan | | |
| Rebecca Wiggins | Trial Manager<br>Centre for Immunology and Infection<br>University of York | | |
| Bachir Atallah | Data Manager<br>ClinServ International<br>Lebanon | | |
| Ada Keding | Statistical Investigator<br>York Trials Unit<br>University of York | | |
| Mohamed Osman | Laboratory Investigator<br>Centre for Immunology and Infection<br>University of York | | |

### 7 Suggested report structure, tables and figures

#### 7.1 Recruitment and Study Progression

Figure 1: Recruitment over time

Figure 2: Patient flow through trial (CONSORT)

Table 1: Reasons for non-completion

#### 7.2 Baseline Characteristics

Table 2: Baseline Characteristics by Allocation

Table 3: Baseline Characteristics by Age Group

#### 7.3 Baseline and Follow-up Measurements

Table 4: Vital Signs, Biochemistry, Haematology by Allocation (Total)

Table 5: Change Vital Signs, Biochemistry, Haematology by Allocation (Total)

Table 6: Vital Signs, Biochemistry, Haematology by Allocation (Adolescents)

Table 7: Change Vital Signs, Biochemistry, Haematology by Allocation (Adolescents)

Table 8: Vital Signs, Biochemistry, Haematology by Allocation (Adults)

Table 9: Change Vital Signs, Biochemistry, Haematology by Allocation (Adults)

#### 7.4 Analysis of Primary Outcomes

##### 7.4.1 Efficacy

Table 10: PKDL Recovery over Time by Allocation

Figure 3: Bar chart of PKDL Recovery over Time by Allocation (≥90% improvement)

Figure 4: Bar chart of PKDL Recovery over Time by Allocation (≥75% improvement)

##### 7.4.2 Safety

Table 10: Summary of Serious Adverse Events by Allocation (Total)

Table 11: Summary of Non-Serious Adverse Events by Allocation (Total)

Figure 4: Box Plot of Number of Serious Adverse Events per Patient by Allocation (Total)

Figure 5: Box Plot of Number of Non-serious Adverse Events per Patient by Allocation (Total)

Table 12: Summary of Serious Adverse Events by Allocation (Adolescents)

Table 13: Summary of Non-Serious Adverse Events by Allocation (Adolescents)

Table 14: Summary of Serious Adverse Events by Allocation (Adults)

Table 15: Summary of Non-Serious Adverse Events by Allocation (Adults)

Table 16: Comparison of Adverse Events per Patient by Allocation

Table 17: Full Listing of all Adverse Events

#### 7.5 Analysis of Secondary Outcomes

Table 18: PKDL Overall Lesions Grade over Time by Allocation (Total)
Table 19: PKDL Lesions Grade by Body Part over Time by Allocation (Total)
Table 20: PKDL Body Part Lesion Sum Score over Time by Allocation (Total)
Table 21: PKDL Overall Lesions Grade over Time by Allocation (Adolescents)
Table 22: PKDL Lesions Grade by Body Part over Time by Allocation (Adolescents)
Table 23: PKDL Body Part Lesion Sum Score over Time by Allocation (Adolescents)
Table 24: PKDL Overall Lesions Grade over Time by Allocation (Adults)
Table 25: PKDL Lesions Grade by Body Part over Time by Allocation (Adults)
Table 26: PKDL Body Part Lesion Sum Score over Time by Allocation (Adults)
